# Individualized treatment effects of corticosteroids in IgA nephropathy

**DOI:** 10.1101/2025.10.08.25337548

**Authors:** D. L. Hölscher, N. E. J. Schmitz, L. Niggemeier, P. Pilva, M. Strauch, V. Tesar, J. Barratt, I. S. D. Roberts, R. Coppo, the VALIGA investigators, L. Barisoni, the CureGN investigators, M. Yanagita, U. Alabalik, A. D. Rule, J. M. Jagtap, E. S. Abreu, M. W. Taal, P. A. Kalra, the NURTuRE academic steering group, J. Floege, R. Kramann, P. Boor, the AI4IgAN study, R. D. Bülow

**Author notes:** ϕ VALIGA, CureGN, NURTuRE & AI4IgAN contributor information are provided in the Supplement. Shared senior authors (P. Boor, R. D. Bülow). Address correspondence to: Peter Boor, M.D., Ph.D., Institute of Pathology, RWTH Aachen University Hospital, Pauwelsstrasse 30, 52074 Aachen, Germany.

## Abstract

IgA nephropathy (IgAN) is a leading cause of kidney failure with diverse clinical presentations and treatment responses, particularly to corticosteroids, with conflicting evidence. Due to potentially severe side effects and heterogenous responses more individualized approaches are needed. We developed a causal machine learning framework for predicting individualized treatment effects of corticosteroids in IgAN by integrating clinical variables, histopathological scores, and deep learning-based biomarkers from digitized kidney biopsies (pathomics) of 1,022 patients from eight retrospective international cohorts. At the cohort-level, corticosteroids showed no significant effect on five-year kidney survival. However, the framework identified subpopulations with and without significant treatment benefit, improving progression-free kidney survival and also reducing overtreatment in low-benefit patients. Pathomics highlighted tubulointerstitial inflammation and glomerular tuft deformation as predictors of corticosteroid response. Our framework offers a blueprint for precision therapy in IgAN, supporting clinical decision-making in the era of emerging targeted treatments.

## Introduction

Immunoglobulin A nephropathy (IgAN) is the most common glomerulonephritis worldwide and a leading cause of kidney failure^1,2^. It remains a complex and diverse entity with highly variable clinical presentations and progression rates^1^, ranging from less than 10% to over 50%^3^ of patients developing kidney failure within 10-20 years of diagnosis. While optimized supportive care (generic chronic kidney disease (CKD) therapy) remains the cornerstone of IgAN management, patients at high risk of progression may benefit from systemic corticosteroids to reduce glomerular inflammation^4,5^. Yet, evidence on corticosteroid efficacy from observational studies and randomized controlled trials is conflicting^6–9^, possibly reflecting insufficient identification of patients most likely to benefit^10^. Such predictive biomarkers are urgently needed, since side-effects of systemic corticosteroid treatment can be life-threatening^7,11,12^.

Clinical trials traditionally report the average effect of a treatment on the defined outcome, which may obscure important heterogeneity in individual responses. Although traditional statistical methods aim to identify subgroups of patients, these one-at-a-time analyses often cannot adequately capture multivariate interactions impacting treatment benefits. Causal machine learning (ML) provides an alternative approach by modeling counterfactual outcomes, which are potential outcomes under alternate treatment scenarios. This enables prediction of individualized treatment effects, i.e., the predicted difference in outcomes between treatment assignments for each individual patient based on their unique characteristics. However, reliable predictors of corticosteroid treatment benefit beyond proteinuria and kidney function have not been determined yet^10^.

Nephropathology is a cornerstone in the diagnostic assessment for IgAN, with the Oxford classification (MEST-C) established as the standard histologic scoring system for patient risk prognostication^13–15^. Although some components of the classification have been linked to corticosteroid treatment benefit^9,16^, current Kidney Disease: Improving Global Outcomes (KDIGO) guidelines do not recommend the use of the Oxford classification to guide treatment decisions due to insufficient supporting evidence^17^. This limitation may be attributable to the semi-quantitative scoring and inter-observer variability encountered in clinical practice^18^.

Gigapixel digital histopathology images, so-called Whole Slide Images (WSIs), contain vast information that can be extracted using deep learning-based models^19^. Pathomics enables automatic quantification of reproducible, explainable, and precise digital biomarkers directly from digitized kidney biopsies in a high-throughput fashion^20^. These digital biomarkers can capture novel information that can be utilized to inform treatment decisions for individual patients in a precision medicine framework.

In this retrospective, worldwide multicenter study of 1,022 patients with biopsy-proven IgAN, we developed and externally validated a causal ML framework that integrates clinical data, histopathological scores, and pathomics to predict individualized treatment effects of systemic corticosteroids. Our approach addresses a critical gap: the need for more personalized treatment decisions that leverage novel data derived from kidney biopsies, in contrast to the current reliance on pooled treatment effects in IgAN. With the emergence of novel therapeutic agents^21–23^ and molecular targets^24^, this framework might serve as a blueprint for advancing patient-tailored treatment strategies in IgAN.

## Results

### Patient characteristics and outcomes

Both the multicentric derivation (n=464, VALIGA, Kyoto, NURTuRE-CKD) and the multicentric validation cohort (n=558, VALIGA, Leicester, Aachen, Diyarbakir, Rochester, CureGN) exhibited similar baseline characteristics and follow-up durations (Figure 1, Table 1, Supplementary Tables 1-2). Notable differences were observed in the histopathological Oxford classification scores, with the validation cohort demonstrating a higher prevalence of mesangial hypercellularity (55.9% vs 31.9%), endocapillary hypercellularity (24.0% vs 16.2%), and presence of crescents (21.0% vs 13.6%), generally indicating more active disease states. Renin-angiotensin system inhibitor (RASi) usage was consistent between cohorts (80.2% vs 80.8%), whereas corticosteroid therapy was more common in the derivation cohort (34.3% vs 20.1%) and administered for a longer duration (1.1 vs 0.6 years). The primary composite outcome (≥ 50% eGFR decline and/or kidney failure within 5 years) occurred in 8.6% (n=40) of the derivation cohort and 8.8% (n=49) of the validation cohort, indicating similar event rates between the two study populations (log-rank test, *p* = 0.79, Supplementary Figure 1A). At the cohort-level, corticosteroids showed no significant improvement in progression-free kidney survival (log-rank test, *p* = 0.27, Supplementary Figure 1B).

**Figure 1.**
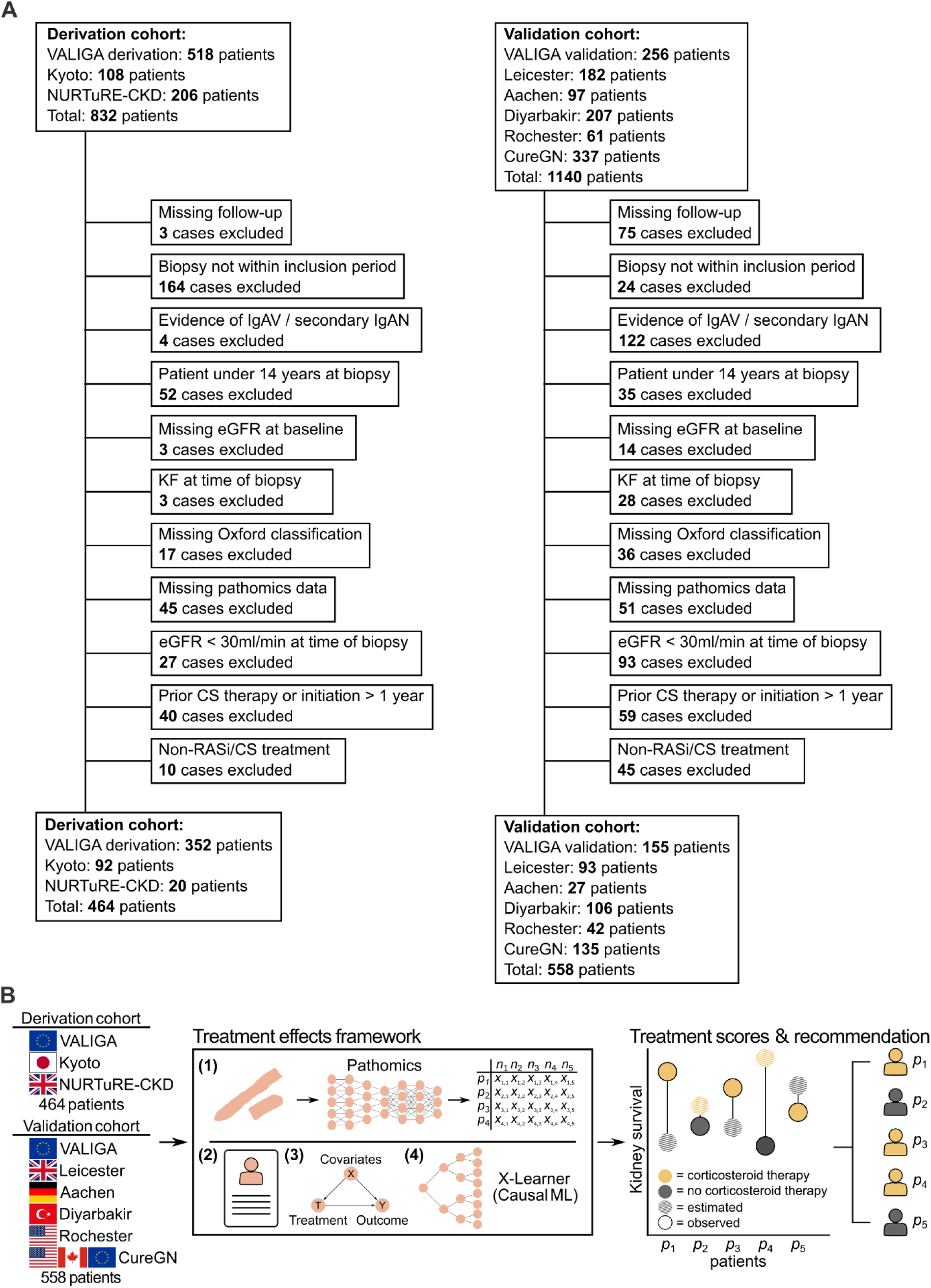
Overview of the proposed framework for individualized treatment effects prediction. Eight cohorts from Europe, North America and Asia were used for derivation and validation with the cohort refinement flowchart shown in (A). (B) All patients from the eight cohorts were analyzed using the causal ML framework, including (1) pathomics-derived predictors obtained via digital image analysis and (2) baseline clinical and histopathological descriptors. (3) Treatment heterogeneity based on included covariates was analyzed using an X-learner algorithm (4) which is a meta-learning approach to estimate counterfactual (unobserved) outcomes for each patient under both treatment and control scenarios. Patient-level predictions of the individualized treatment effect are computed, informing the development of a personalized treatment recommendation. Abbreviations: VALIGA, European Validation Study of the Oxford Classification of IgA Nephropathy; NURTuRE-CKD, National Unified Renal Translational Research Enterprise-Chronic Kidney Disease; CureGN, Cure Glomerulonephropathy; IgAV, IgA vasculitis; IgAN, IgA nephropathy; eGFR, estimated glomerular filtration rate; KF, kidney failure; CS, corticosteroid; RASi, renin-angiotensin-system inhibitor; ML, machine learning.

**Table 1.** Characteristics of the derivation and validation cohort. Continuous variables are reported as median (interquartile range) while categorical variables are reported as absolute (n) and relative (%) frequencies. The primary outcome was defined as a composite of ≥ 50% eGFR reduction and kidney failure within five years of kidney biopsy. Abbreviations: m, male; f, female; eGFR, estimated glomerular filtration rate; map, mean arterial blood pressure; M, mesangial hypercellularity; E, endocapillary hypercellularity; S, segmental glomerulosclerosis; T, tubular atrophy and interstitial fibrosis; C, crescents; y, yes; n, no; RASi, renin-angiotensin-system inhibitor; CS, corticosteroid.

|  | Derivation | Validation |
| --- | --- | --- |
| n | 464 | 558 |
| Follow-up [years] | 4.6 (6.1) | 5.0 (6.0) |
| Age [years] | 35.9 (21.1) | 36.7 (20.9) |
| Sex [m\|f] | m: 316 (68.1%) \| f: 148 (31.9%) | m: 364 (65.2%) \| f: 194 (34.8%) |
| eGFR [ml/min/1.73m<sup>2</sup>] | 82.6 (50.1) | 76.8 (52.1) |
| Proteinuria [g/24h] | 1.0 (1.8) | 1.1 (1.6) |
| MAP [mmHg] | 96.7 (16.7) | 93.8 (17.3) |
| M [0\|1] | 0: 316 (68.1%) \| 1: 148 (31.9%) | 0: 246 (44.1%) \| 1: 312 (55.9%) |
| E [0\|1] | 0: 389 (83.8%) \| 1: 75 (16.2%) | 0: 424 (76.0%) \| 1: 134 (24.0%) |
| S [0\|1] | 0: 98 (21.1%) \| 1: 366 (78.9%) | 0: 190 (34.1%) \| 1: 368 (65.9%) |
| T [0\|1\|2] | 0: 369 (79.5%) \| 1: 87 (18.8%) \| 2: 8 (1.7%) | 0: 383 (68.6%) \| 1: 164 (29.4%) \| 2: 11 (2.0%) |
| C [0\|1] | 0: 401 (86.4%) \| 1: 63 (13.6%) | 0: 441 (79.0%) \| 1: 117 (21.0%) |
| Tuft circularity | 0.36 (0.09) | 0.36 (0.1) |
| Tuft eccentricity | 0.68 (0.09) | 0.67 (0.1) |
| Tuft area [$\mu\text{m}^2$] | 6207.01 (8548.22) | 5652.7 (9686.48) |
| Tubular diameter [$\mu\text{m}$] | 27.73 (6.73) | 28.6 (6.75) |
| Tubular distance [$\mu\text{m}$] | 2.13 (1.03) | 2.26 (1.14) |
| RASi [y/n] | y: 372 (80.2%) \| n: 92 (19.8%) | y: 451 (80.8%) \| n: 107 (19.2%) |
| Corticosteroids [y/n] | y: 159 (34.3%) \| n: 305 (65.7%) | y: 112 (20.1%) \| n: 446 (79.9%) |
| CS therapy duration<br>[years] | 1.1 (1.1) | 0.6 (0.6) |
| Outcome [y/n] | y: 40 (8.6%) \| n: 424 (91.4%) | y: 49 (8.8%) \| n: 509 (91.2%) |

### Individualized treatment effects

Individualized treatment effects computed by the causal ML model revealed substantial heterogeneity in predicted treatment benefit (Figure 2B-C). Linear regression analysis demonstrated that a patients’ tertile of predicted benefit (low, middle, high; Figure 2C) significantly modified the observed effect of corticosteroid therapy on the primary composite outcome (likelihood-ratio test for effect modification, *p* < 0.05 in the validation cohort).

**Figure 2.**
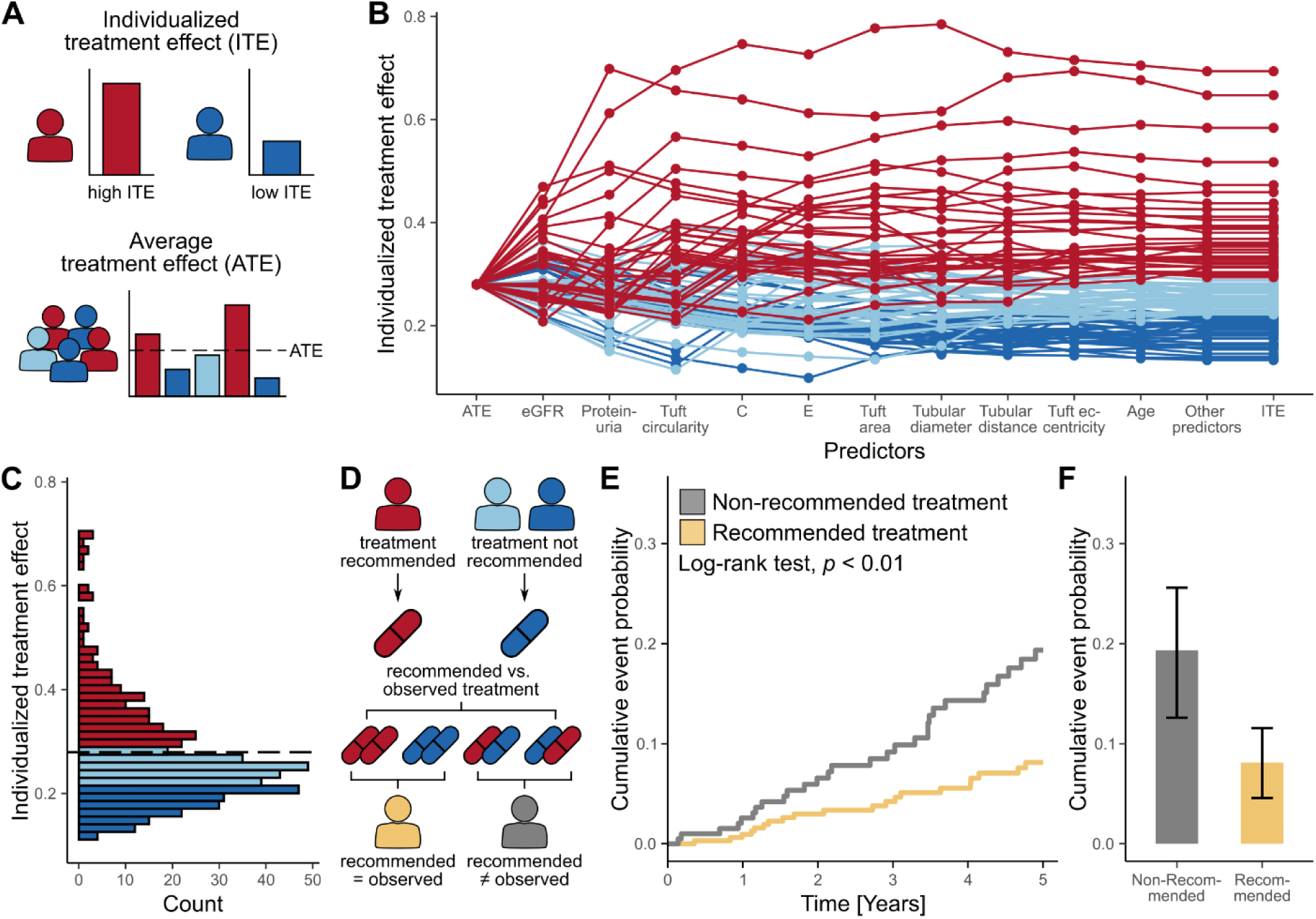
Predicted individualized treatment effects and individual treatment recommendations. (A) Illustration of average treatment effect and individualized treatment effect. (B) Line plot showing the change from the cohort-level average treatment effect to predicted patient-based individualized treatment effects based on the sequential addition of the most-important predictors. Each line represents one patient randomly chosen from each percentile from the 95% distribution of the predicted individualized treatment effect in the validation cohort (C), with patients overall grouped into tertiles based on their predicted benefit (red: upper tertile, high predicted benefit; light blue: middle tertile, medium predicted benefit; blue, lower tertile, low predicted benefit). (D) Implementation of a treatment recommendation only recommending treatment for the individuals with highest predicted benefit (red) - individualized treatment recommendations were then retrospectively compared to the observed treatment (corticosteroid therapy vs. no corticosteroid therapy) during follow-up. (E) Kaplan-Meier curves comparing patients in the validation cohort who received the recommended treatment versus those who did not. (F) Cumulative event probability at five years of both groups including 95% confidence intervals. Abbreviations: eGFR, estimated glomerular filtration rate; C, crescents; E, endocapillary hypercellularity.

Based on the observed heterogeneity of treatment effects, we evaluated an individualized treatment recommendation. Considering the distribution of predicted benefit and the associated uplift gain (Supplementary Figure 2A), we assessed a model-based treatment recommendation targeting patients in the upper tertile, i.e., those with high predicted benefit of corticosteroid treatment. Patients in the middle and lower tertiles were not recommended to receive corticosteroid therapy. We retrospectively compared all model-based treatment recommendations to the observed treatment (Figure 2D). Among patients for whom the model recommended corticosteroid therapy, those who received corticosteroids experienced significantly longer progression-free kidney survival than those who did not (derivation: 0.56 years [95% CI 0.23-0.89], *p* < 0.01; validation: 0.4 years [95% CI 0.13-0.67], *p* < 0.05, Supplementary Figure 3A-B, Supplementary Table 3). In contrast, among patients for whom corticosteroid therapy was not recommended by the model, no significant effect of corticosteroid use was observed (derivation: *p* = 0.2, validation: *p* = 1.0, Supplementary Figure 3A-B, Supplementary Table 3). Implementing these model-based treatment recommendations would have reduced corticosteroid use by 67.3% and 60.7% for patients in the middle and lower tertiles of the derivation and validation cohorts, respectively.

Comparing patients who received the model-recommended treatment to those who did not revealed significantly longer progression-free kidney survival by 0.43 years (95% CI 0.26-0.6, *p* < 0.01, Supplementary Figure 3C) in the derivation and by 0.24 years (95% CI 0.06-0.42, *p* < 0.01, Figure 2E) in the validation cohort. This corresponded to an absolute risk reduction of the composite endpoint at five years of 18.0% in the derivation (Supplementary Figure 3D) and 11.2% in the validation cohort (Figure 2F). Detailed clinical characteristics of all subgroups in both cohorts are presented in Supplementary Tables 4-5.

### Model performance

The uplift in the Qini curve and increase in the rank-weighted average treatment effect (RATE) curve demonstrated a positive association between higher predicted individualized treatment effect and improved progression-free kidney survival (Supplementary Figure 2). This indicates that the model effectively stratified patients according to their expected benefit from corticosteroid treatment, identifying those most likely to experience slower disease progression when treated. The Qini coefficient, i.e., the additional treatment benefit gained by targeting corticosteroid therapy using model predictions, was 7.41 (95% CI 2.57-12.9) in the derivation and 7.08 (95% CI 2.38-13.37) in the validation cohort, with higher values reflecting the improvements in patient outcomes due to personalized treatment allocation. Corresponding RATE was 0.35 (95% CI 0.05-0.66) and 0.28 (95% CI 0.1-0.47), respectively. The model’s ability to discriminate patients with greater or lesser treatment benefit was further supported by a C-for-benefit of 0.63 (95% CI 0.58-0.68) in the derivation and 0.55 (95% CI 0.5-0.6) in the validation cohort.

Overall, these performance metrics confirm the capability of the model to effectively discriminate treatment effects. The model was well calibrated for patients with high predicted benefit, whereas predictions for low-benefit groups were overestimated (Supplementary Figure 4).

### Treatment-covariate interactions

Deconstructing model predictions by Shapley additive explanations (SHAP), identified the most influential predictors determining corticosteroid treatment benefit (Figure 3A), including baseline eGFR, proteinuria, glomerular tuft circularity, presence of crescents (C1), endocapillary hypercellularity (E1), and tuft area. Lower baseline eGFR and higher proteinuria were most strongly associated with increased treatment benefit (Figure 3B+C), followed by pathomics and MEST-C scores. The relative influence of the ten most important predictors on patients’ predicted treatment effect is illustrated in Figure 2B, and representative cases of high and low predicted benefit are depicted in Supplementary Figure 5.

**Figure 3.**
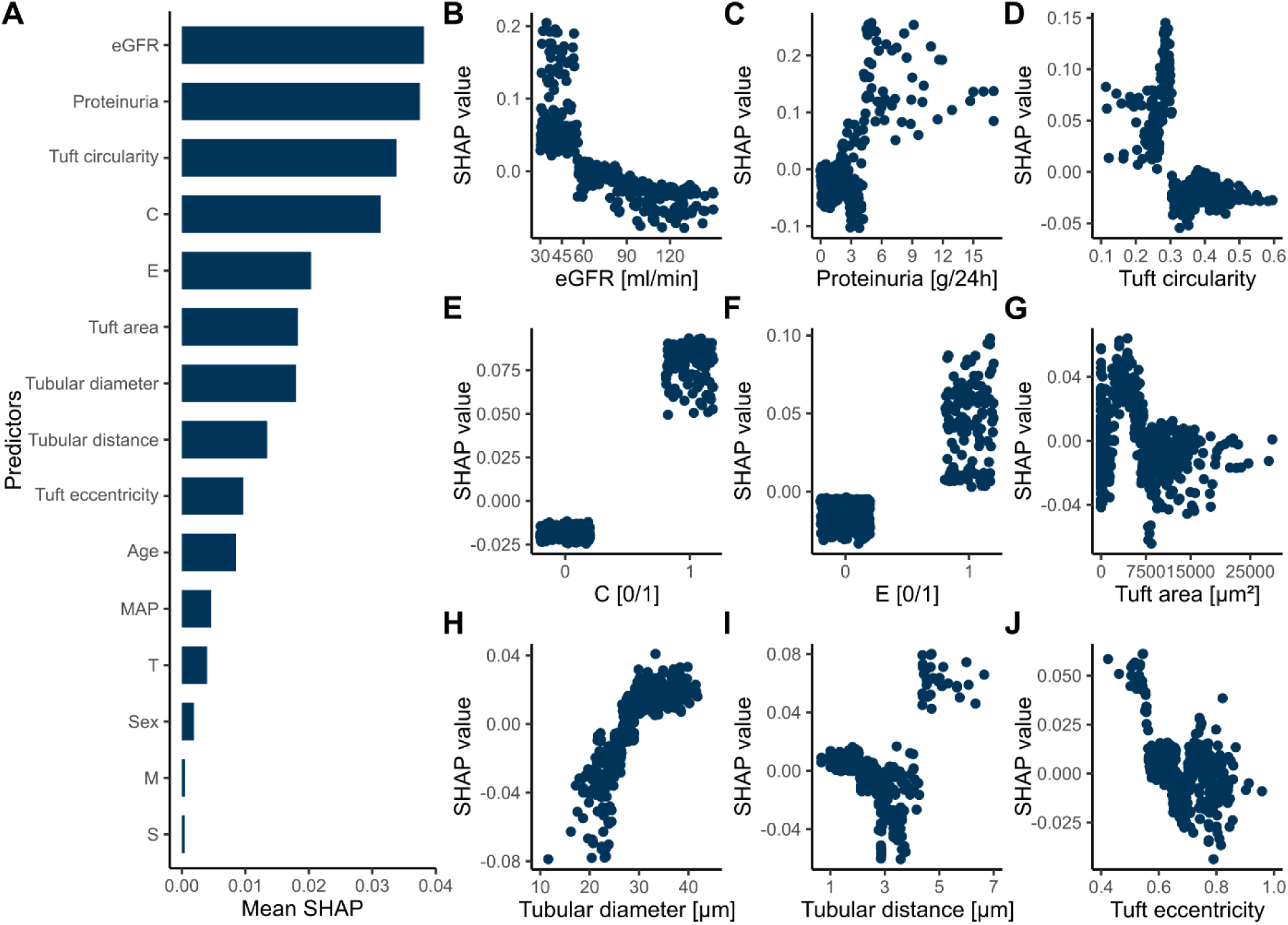
Derived Shapley additive explanations (SHAP) from the validation cohort. (A) Aggregation of global feature importance and influence of the nine most important clinical, histopathological, and pathomics-based predictors on the individualized treatment effect on patient-level (B-J). Higher SHAP values correspond to a greater predicted treatment benefit of corticosteroid therapy. Abbreviations: eGFR, estimated glomerular filtration rate; C, crescents; E, endocapillary hypercellularity.

SHAP values of the Oxford classification components demonstrated the importance of active lesions (E1/C1) as a sign of glomerular inflammation. In contrast, progressive tubulointerstitial scarring (T1/T2) was associated with less benefit. For mesangial hypercellularity (M1) and segmental glomerulosclerosis (S1) no marked effect was detected (Supplementary Figure 6).

Among pathomics-derived predictors, deformation of the glomerular tuft, marked by lower tuft circularity (irregular shape) and tuft eccentricity (globular shape), was associated with greater treatment benefit (examples of glomeruli based on tuft circularity and predicted benefit are shown in Supplementary Figure 7). The tuft area exhibited a parabolic relationship with decreased benefit at the tails of the distribution, likely corresponding to sclerotic glomeruli with a small tuft area and hypertrophic glomeruli with a large tuft area (Figure 3G). Tubular distance and diameter also ranked among the ten most influential predictors (Figure 3A), with smaller tubular diameter and greater tubular distance (indicative of tubular atrophy and interstitial fibrosis) generally associated with decreased treatment benefit (Figure 3H+I). However, a subset of patients with markedly increased tubular distance was unexpectedly shown to benefit from corticosteroid therapy. Partial dependence analysis revealed a specific combination of high tubular distance and high tubular diameter associated with high predicted benefit (Figure 4A). Histopathological assessment comparing cases with this pathomics pattern (n=30) versus those with high tubular distance and low predicted benefit (n=30) identified a significant increase in interstitial inflammation (*p* < 0.05) and tubulitis (*p* < 0.05). Patients exhibiting this distinct pathomics pattern were four times more likely to exhibit interstitial inflammation (i1-3, Figure 4B) and 2.8 times more likely to show tubulitis (t1-3, Figure 4C, Supplementary Table 6). Representative images of these cases are provided in Figure 4F+G.

**Figure 4.**
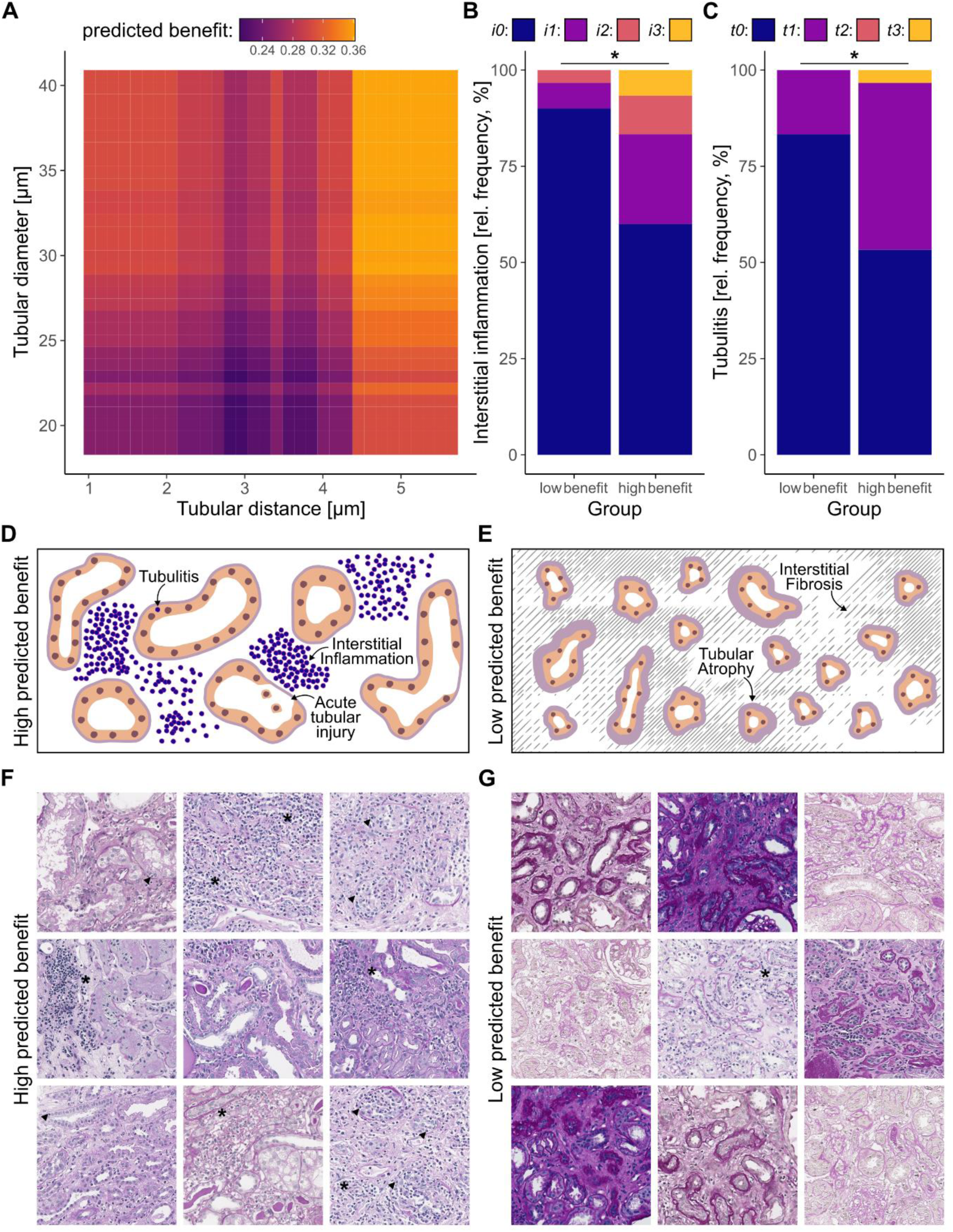
Identification of a tubulointerstitial pathomics pattern representing tubulointerstitial inflammation. (A) Partial dependence plot of tubular distance and tubular diameter depicting the combined effect of tubulointerstitial pathomics features on the predicted treatment benefit. Pathologist scoring demonstrated that interstitial inflammation (i, B) and tubulitis (t, C) occurred more often in representative patients with higher predicted treatment benefit compared to those with lower benefit. Patients with high predicted benefit displayed more acute inflammatory IgAN disease activity (D, F) and acute tubular injury, while patients with low predicted benefit exhibited more chronic changes (E, G) such as tubular atrophy and interstitial fibrosis. Arrows show tubulitis while stars highlight areas with increased interstitial inflammation. All images represent patches of 300x300µm.

### Sensitivity and robustness

To validate the model predictions, we conducted multiple sensitivity and robustness analyses. We further analyzed potential cohort-specific effects by including an interaction term between corticosteroid treatment and patient cohort to test for effect modification. The likelihood-ratio tests for effect modification showed no significant heterogeneity in observed (*p* = 0.38) or predicted (*p* = 0.21) treatment effects across cohorts, indicating that corticosteroid benefits were consistent and not dependent on cohort-specific factors (Supplementary Figure 8). Additionally, we found no significant modification of the observed or predicted treatment effect by race and ethnicity, although our analysis was underpowered, especially for Chinese patient populations (likelihood-ratio test for effect modification, *p* = 0.34 and *p* = 0.31). Interestingly, the duration of corticosteroid treatment (shorter treatment duration of up to 6 months vs. longer treatment duration of over 6 months) did not significantly modify the observed (*p* = 0.33) or predicted treatment (*p* = 0.78) effect in treated patients.

Robustness analysis of introducing a random predictor did not substantially alter overall model performance, as the predicted average treatment effect remained consistent. In contrast, replacing proteinuria with a random continuous variable led to a notable difference in predicted average treatment effect. Moreover, randomly assigning treatment exposure or outcomes resulted in a marked decline in predicted average treatment effect, confirming the model’s reliance on biologically meaningful relationships in the data (Supplementary Table 7).

## Discussion

In this retrospective, worldwide multicenter study, we developed and validated a causal ML model for the prediction of individualized treatment effects of systemic corticosteroid therapy in IgAN patients. In line with previous trials on the effectiveness of corticosteroid treatment^6,25,26^, we could not observe a pooled significant benefit of systemic corticosteroids in our study. In contrast, utilizing the causal ML framework we identified a subset of patients who exhibited a significant clinical benefit, with an absolute risk reduction of the primary composite outcome of 11.2% at five years in the validation cohort. By specifically identifying patients with a high likelihood of benefiting from systemic corticosteroid therapy, usage of our model would have theoretically reduced unnecessary corticosteroid exposure by 60.7% in the validation cohort, while at the same time improving patient outcomes.

The key predictors identified by our model provide plausibility for treatment response predictions: lower eGFR, higher proteinuria, and specific Oxford classification scores (C1, E1), potentially reflecting inflammatory disease activity. Presence of crescents (C1-Score) and endocapillary hypercellularity (E1-Score) have previously been associated with response to immunosuppressive therapy^27,28^. Patients with lower tuft circularity, a morphometric pattern suggesting glomerular tuft distortion, showed greater benefit from corticosteroid therapy. We have previously shown that this feature is indicative of a more severe disease state in IgAN^29^. Additionally, pathomics predictors reflecting tubulointerstitial changes could identified a subset of patients with tubulointerstitial inflammation. This lesion was initially omitted from the Oxford classification due to poor reproducibility^13^, yet multiple studies indicate its potential role in disease progression^30,31^, although results on corticosteroid effectiveness were underpowered^31^. In our study, pathomics enabled a precise characterization of the tubulointerstitial architecture, an aspect whose importance has also been previously demonstrated in other glomerular diseases^32^. Whether tubulointerstitial inflammation reflects IgAN disease activity or represents a separate overlapping condition benefiting from corticosteroid treatment remains to be further investigated. Nonetheless, reporting the degree of tubulointerstitial inflammation by pathologists could be easily implemented into current practice, e.g., by reporting the widely used Banff lesion scores *i* and *t* in IgAN biopsies.

By identifying patient-specific baseline characteristics as modifiers of corticosteroid treatment effect, our approach provides a potential explanation as to why previous pooled analyses have failed to demonstrate consistent benefits. This paradigm shift from generic to individualized treatment decisions addresses a fundamental constraint in current IgA nephropathy management.

Our study has some limitations. First, the retrospective design carries the potential for unmeasured confounding factors. Prospective validation in randomized controlled trials will be essential to confirm the clinical utility of model-based treatment decisions. Second, differences in data collection standards, reporting of predictors, and treatment interventions could limit the predictive performance of our model, although a sensitivity analysis did not show any substantial influence of cohort heterogeneity. Importantly, dosing of systemic corticosteroid and RASi medication was reported inconsistently. Third, our analysis is limited to initiation of therapy within a year after biopsy, thus, we cannot account for any longitudinal changes that might inform treatment decisions after the specified timeframe. Also, given the specified follow-up period of 5 years, the long-term effects of treatment on the trajectory of disease progression remain to be investigated, potentially partly explaining the only modest improvement of progression-free survival in the high benefit group and the importance of kidney function at time of biopsy. Our current analysis focused on systemic corticosteroids, but the developed framework could be extended to other immunosuppressive therapies, novel treatments and integration with emerging biomarkers and complement activation markers in the future.

Our findings reveal that apparent treatment failures at the population-level may mask significant benefits in well-defined patient subgroups, highlighting the importance of accounting for treatment effect heterogeneity in clinical research and practice. Using highly granular pathomics features might aid this effort. Our multimodal approach could be readily adapted to the rapidly expanding treatment landscape of IgAN as well as other glomerular diseases, where tissue-based biomarkers can guide individualized decision-making and precision nephrology.

## Methods

### Study population

This retrospective analysis consists of data from eight international cohorts comprising a total of 1,022 patients with biopsy-proven IgA nephropathy which were used for model derivation and validation. The study cohorts include a secondary analysis of corticosteroids in the *European Validation Study of the Oxford Classification of IgA Nephropathy* (VALIGA) cohort^14^, a European multi-center cohort, two multi-center repositories (*Cure Glomerulonephropathy* (CureGN), North America and Europe) and a multi-center national CKD cohort (*National Unified Renal Translational Research Enterprise* (NURTuRE-CKD, United Kingdom^33^) as well as several single-center cohorts from Kyoto (Japan), Leicester (United Kingdom), Diyarbakir (Turkey), Rochester (United States of America) and Aachen (Germany). A detailed overview of the cohort refinement process and relevant exclusion criteria is provided in Figure 1A and the Supplementary Methods. The VALIGA cohort was split by clinical center into derivation and validation subsets to achieve better balance in covariates, treatment regimens and outcomes across the cohorts^34^. In total, the refined derivation cohort comprised 464 patients from the VALIGA, Kyoto and NURTuRE-CKD cohorts and the validation cohort consisted of 558 patients from the VALIGA, Leicester, Aachen, Diyarbakir, Rochester and CureGN cohorts.

### Study design

This study applied an effect-based analysis of heterogeneity of treatment effect (HTE), as outlined in the effect-modelling framework of the *Predictive Approaches to Treatment Effect Heterogeneity* (PATH) statement^35^. Reporting of study populations, outcomes, predictors, model development, and validation adhered to the *Transparent Reporting of a Multivariable Prediction Model for Individual Prognosis or Diagnosis* (TRIPOD+AI) guideline^36^ (Supplementary Table 8). All patients were assessed at time of kidney biopsy with subsequent follow-up for a period of five years. Treatment was defined as initiation of systemic corticosteroid therapy within the first year after biopsy. Patients receiving other immunosuppressants, targeted-release corticosteroid formulations or dual endothelin and angiotensin II receptor antagonists (DERAs) were excluded. The primary composite outcome was progression of IgA nephropathy, defined as a sustained ≥ 50% decline in eGFR and/or development of kidney failure, including persistent eGFR below 15ml/min/1.73m², initiation of dialysis or kidney transplantation. Outcomes were analyzed using the restricted mean survival time (RMST) in years after kidney biopsy.

### Predictors

For all included patients demographic and clinical parameters including age, sex, race and ethnicity, body-mass-index (BMI), eGFR, proteinuria, systolic, diastolic and mean arterial blood pressure (MAP) were recorded at time of biopsy. In addition, all biopsies were scored according to the Oxford classification MEST-C scoring system^15,37^. We further analyzed five biopsy-level pathomics predictors of glomerular (glomerular tuft area, tuft circularity and tuft eccentricity) and tubulointerstitial morphometry (tubular diameter and tubular distance) for each patient (Supplementary Table 9). These five pathomics predictors were previously demonstrated to be associated with long-term outcomes in IgAN^29^.

### Statistical analysis

We employed an Xboost model to estimate heterogeneous treatment effects due to its advantages for unbalanced treatment assignment and suitability for retrospective observational studies^38^. Xboost is an XGBoost-based implementation of the X-learner, a meta-learning framework for causal inference^39^. XGBoost is a tree-based, non-parametric machine learning algorithm that incorporates advanced regularization to reduce overfitting^40,41^. The X-learner framework consists of two primary stages including computing of pseudo-outcomes followed by propensity score modeling to adjust for confounding^38^. Further details on the framework are included in the Supplementary Methods (Model architecture). Hyperparameter tuning was conducted, and robust model performance was ensured using 10-fold cross-validation in the derivation cohort. The final fitted model was subsequently applied to the validation cohort. Model performance was assessed in both cohorts using the Qini coefficient, rank-weighted average treatment effect (RATE) and C-for-benefit (see Supplementary Methods, Model performance). Model calibration was evaluated in the validation cohort by comparing predicted versus observed treatment effects.

To illustrate treatment heterogeneity identified by the model, patients were ranked based on their predicted individualized treatment effect, i.e., the difference in RMST with versus without corticosteroid therapy, and then stratified into tertiles in accordance with the PATH statement^35^. We then implemented a treatment recommendation for corticosteroid therapy based on the Qini curve and distribution of individualized treatment effect in comparison to the overall average treatment effect. Patients in the upper third of predicted individualized treatment effect with high expected benefit of corticosteroid therapy were categorized as recommended to be treated with corticosteroids while the middle and lower tertile with low expected benefit were recommended not to be treated. The interaction between baseline risk and treatment benefit in these groups was assessed by linear regression and likelihood ratio testing. To further assess clinical utility, we evaluated patient outcomes and their RMST comparing the respective treatment recommendation and the observed treatment with Bonferroni-type adjustment for multiple testing.

We evaluated the relative contribution of each baseline characteristic in predicting individualized treatment effects by implementing Shapley Additive Explanations (SHAP) and partial dependence analyses on model predictions in the validation cohort.

Furthermore, we performed multiple sensitivity and robustness analyses to validate the predictions of our model. Full details of these post-hoc analyses are provided in the Supplementary Methods (Sensitivity and robustness analyses).

All variables, including clinical and pathomics predictors, were summarized by cohort and subgroup, with continuous variables reported as median and interquartile range (IQR) and categorical variables as absolute (n) and relative (%) frequencies. All statistical tests were two-sided, with significance defined as *p* < 0.05. All calculations were performed within R version 4.4.3.

## Data and code availability

Whole slide image files and clinical data used in this study are available under restricted access for privacy protection reasons. Access can be obtained by directly contacting the corresponding author who can distribute requests to the collaborators involved who contributed data. Alternatively, to obtain access for individual datasets, all included study groups and contributing collaborators are provided in Supplementary Table 10. In general, requests will be evaluated within a timeframe of 4 weeks based on institutional and trial policies.

Annotated code on model derivation and validation as well as all subsequently performed analyses is available under GitLab (https://git-ce.rwth-aachen.de/labooratory-ai/igan-ite-prediction).

## Ethics statement

Data collection and analysis in this study was performed in accordance with the Declaration of Helsinki and was approved by the local ethics committee of the RWTH Aachen University (EK-No. 125/25).

## Acknowledgments

Funding for the CureGN consortium is provided by U24DK100845, U01DK100846, U01DK100876, U01DK100866, and U01DK100867 from the National Institute of Diabetes and Digestive and Kidney Diseases (NIDDK). Patient recruitment is supported by NephCure. The views expressed in written materials or publications do not necessarily reflect the official policies of the Department of Health and Human Services; nor does mention by trade names, commercial practices, or organizations imply endorsement by the U.S. Government.

The authors gratefully acknowledge the contributions of all colleagues and collaborators from the VALIGA, CureGN, NURTuRE and AI4IgAN groups. We also would like to thank all patients who participated in this study.

## Author contributions

Conceptualization: DLH, RDB; Methodology: DLH; RDB; Software: DLH, NEJS, LN, PP, MS, JMJ; Validation: DLH, RDB; Formal analysis: DLH; Investigation: DLH; Resources: PB, RDB; Data Curation: DLH, NEJS, LN, PP, MS, VT, JB, ISDR, RC, LB, MY, UA, ADR, ESA, MWT, PAK, JF, RK, PB, RDB; Writing - Original Draft: DLH, RDB; Writing - Review & Editing: DLH, NEJS, PP, MS VT, JB, ISDR, RC, LB, MY, UA, ADR, JMJ, ESA, MWT, PAK, JF, RK, PB, RDB; Visualization: DLH; Supervision: RDB, PB; Project administration: None; Funding acquisition: PB, RDB.

