## Supplementary Material for "Individualized treatment effects of corticosteroids in IgA nephropathy"

### Supplementary Material to: Individualized treatment effects of corticosteroids in IgA nephropathy

### Table of Contents

Group Information

Supplementary Methods

- Cohort refinement
- Definitions for predictors, outcomes and treatments
- Computation of pathomics features
- Model architecture
- Model assessment
- Partitioning of cohorts and treatment policy evaluation
- Sensitivity and robustness analyses

Supp. Table 1: Additional baseline characteristics

Supp. Table 2: Baseline patient characteristics in the individual cohorts

Supp. Table 3: Restricted mean progression-free survival based on treatment policy

Supp. Table 4: Derivation cohort: characteristics of partitioned subgroups

Supp. Table 5: Validation cohort: characteristics of partitioned subgroups

Supp. Table 6: Scoring of tubulointerstitial inflammation

Supp. Table 7: Robustness analyses

Supp. Table 8: TRIPOD+AI checklist

Supp. Table 9: Definitions for pathomics features

Supp. Table 10: Overview of datasets, study groups and contributors

Supp. Table 11: Missingness of predictors in derivation and validation cohort

Supp. Figure 1: Baseline cumulative event probability and overall treatment effect

Supp. Figure 2: Qini and rank-weighted average treatment effect

Supp. Figure 3: Cumulative event probability for treatment recommendations

Supp. Figure 4: Model calibration

Supp. Figure 5: SHAP values of representative cases

Supp. Figure 6: SHAP values of MEST-C predictors

Supp. Figure 7: Representative visualizations of glomeruli in patients with high and low predicted treatment benefit

Supp. Figure 8: Distribution of ITE within cohorts and subcohorts

### Group Information

**The VALIGA & International IgA Nephropathy Network members are as follows:**
VALIGA investigators: M.L. Russo (MA, PhD, Fondazione Ricerca Molinette, Torino, Italy); S. Troyanov (MD, Division of Nephrology, Department of Medicine, Hopital du Sacre-Coeur de Montreal, Montreal, Quebec, Canada); H.T. Cook (MD, Centre for Complement and Inflammation Research, Department of Medicine, Imperial College, London, England); I.S.D. Roberts (MD, Department of Cellular Pathology, Oxford University Hospitals NHS Foundation Trust, John Radcliffe Hospital, Oxford, United Kingdom); V. Tesar (MD, Department of Nephrology, 1st Faculty of Medicine and General University Hospital, Charles University, Prague, Czech Republic); D. Maixnerova (MD, Department of Nephrology, 1st Faculty of Medicine and General University Hospital, Charles University, Prague, Czech Republic); S. Lundberg (MD, Nephrology Unit, Department of Clinical Sciences, Karolinska Institute, Stockholm, Sweden); L. Gesualdo (MD, Department of Nephrology, Emergency and Organ Transplantation, University of Bari "Aldo Moro," Foggia-Bari, Italy); F. Emma (MD, Division of Nephrology, Department of Pediatric Subspecialties, Bambino Gesù Children's Hospital IRCCS, Rome, Italy); F. Diomedi (MD, Division of Nephrology, Department of Pediatric Subspecialties, Bambino Gesù Children's Hospital IRCCS, Rome, Italy); G. Beltrame (MD, Nephrology and Dialysis Unit, San Giovanni Bosco Hospital, and University of Turin, Turin, Italy); C. Rollino (MD, Nephrology and Dialysis Unit, San Giovanni Bosco Hospital, and University of Turin, Turin, Italy); A. Amore (MD, Nephrology Unit, Regina Margherita Children's Hospital, Turin, Italy); R. Camilla (MD Nephrology Unit, Regina Margherita Children's Hospital, Turin, Italy); L. Peruzzi (MD, Nephrology Unit, Regina Margherita Children's Hospital, Turin, Italy); M. Praga (MD, Nephrology Unit, Hospital 12 de Octubre, Madrid, Spain); S. Feriozzi (MD, Nephrology Unit, Belcolle Hospital, Viterbo, Italy), R. Polci, (MD, Nephrology Unit, Belcolle Hospital, Viterbo, Italy); G. Segoloni, (MD, Division of Nephrology Dialysis and Transplantation, Department of Medical Sciences, Città della Salute e della Scienza Hospital and University of Turin, Turin, Italy); L. Colla (MD, Division of Nephrology Dialysis and Transplantation, Department of Medical Sciences, Città della Salute e della Scienza Hospital and University of Turin, Turin, Italy); A. Pani (MD, Nephrology Unit, G. Brotzu Hospital, Cagliari, Italy); D. Piras (MD, Nephrology Unit, G. Brotzu Hospital, Cagliari, Italy), A. Angioi (MD, Nephrology Unit, G. Brotzu Hospital, Cagliari, Italy); G. Cancarini, (MD, Nephrology Unit, Spedali Civili University Hospital, Brescia, Italy); S. Ravera (MD, Nephrology Unit, Spedali Civili University Hospital, Brescia, Italy); M. Durlik (MD, Department of Transplantation Medicine, Nephrology, and Internal Medicine, Medical University of Warsaw, Warsaw, Poland); E. Moggia (Nephrology Unit, Santa Croce Hospital, Cuneo, Italy); J. Ballarin (MD, Department of Nephrology, Fundacion Puigvert, Barcelona, Spain); S. Di Giulio (MD, Nephrology Unit, San Camillo Forlanini Hospital, Rome, Italy); F. Pugliese (MD, Department of Nephrology, Policlinico Umberto I University Hospital, Rome, Italy); I. Serriello (MD, Department of Nephrology, Policlinico Umberto I

University Hospital, Rome, Italy); Y. Caliskan (MD, Division of Nephrology, Department of Internal Medicine, Istanbul Faculty of Medicine, Istanbul University, Istanbul, Turkey); M. Sever (MD, Division of Nephrology, Department of Internal Medicine, Istanbul Faculty of Medicine, Istanbul University, Istanbul, Turkey); I. Kilicaslan (MD, Department of Pathology, Istanbul Faculty of Medicine, Istanbul University, Istanbul, Turkey); F. Locatelli (MD, Department of Nephrology and Dialysis, Alessandro Manzoni Hospital, ASST Lecco, Italy); L. Del Vecchio (MD, Department of Nephrology and Dialysis, Alessandro Manzoni Hospital, ASST Lecco, Italy); J.F.M. Wetzels (MD, Departments of Nephrology, Radboud University Medical Center, Nijmegen, the Netherlands); H. Peters (MD, Departments of Nephrology, Radboud University Medical Center, Nijmegen, the Netherlands); U. Berg (MD, Division of Pediatrics, Department of Clinical Science, Intervention and Technology, Huddinge, Sweden); F. Carvalho (MD, Nephrology Unit, Hospital de Curry Cabral, Lisbon, Portugal); A.C. da Costa Ferreira (MD, Nephrology Unit, Hospital de Curry Cabral, Lisbon, Portugal); M. Maggio (MD, Nephrology Unit, Hospital Maggiore di Lodi, Lodi, Italy); A. Wiecek (MD, Department Nephrology, Endocrinology and Metabolic Diseases, Silesian University of Medicine, Katowice, Poland); M. Ots-Rosenberg (MD, Nephrology Unit, Tartu University Clinics, Tartu, Estonia); R. Magistroni (MD, Department of Nephrology, Policlinic of Modena and Reggio Emilia; Modena, Italy); R. Topaloglu (MD, Department of Pediatric Nephrology and Rheumatology, Hacettepe University, Ankara, Turkey); Y. Bilginer (MD, Department of Pediatric Nephrology and Rheumatology, Hacettepe University, Ankara, Turkey); M. D'Amico (MD, Nephrology Unit, S. Anna Hospital, Como, Italy); M. Stangou (MD, Department of Nephrology, Hippokration General Hospital, Aristotle University of Thessaloniki, Thessaloniki, Greece); F. Giacchino (MD, Nephrology Unit, Ivrea Hospital, Ivrea, Italy); D. Goumenos (MD, Department of Nephrology, University Hospital of Patras, Patras, Greece); P. Kalliakmani (MD, Department of Nephrology, University Hospital of Patras, Patras, Greece); M. Papasotiriou (MD Department of Nephrology, University Hospital of Patras, Patras, Greece); K. Galesic (MD, Department of Nephrology, University Hospital Dubrava, Zagreb, Croatia); C. Geddes (MD, Renal Unit, Western Infirmary Glasgow, Glasgow, United Kingdom); K. Siamopoulos (MD, Nephrology Unit, Medical School University of Ioanina, Ioannina, Greece); O. Balafa (MD, Nephrology Unit, Medical School University of Ioanina, Ioannina, Greece); M. Galliani (MD, Nephrology Unit, S.Pertini Hospital, Rome, Italy); P. Stratta (MD, Department of Nephrology, Maggiore della Carità Hospital, Piemonte Orientale University, Novara, Italy); M. Quaglia (MD, Department of Nephrology, Maggiore della Carità Hospital, Piemonte Orientale University, Novara, Italy); R. Bergia (MD, Nephrology Unit, Degli Infermi Hospital, Biella, Italy); R. Cravero (MD, Nephrology Unit, Degli Infermi Hospital, Biella, Italy); M. Salvadori, (MD, Department of Nephrology, Careggi Hospital, Florence, Italy); L. Cirami (MD, Department of Nephrology, Careggi Hospital, Florence, Italy); B. Fellstrom (MD, Renal Department, University of Uppsala, Uppsala, Sweden); H. Kloster Smerud (MD, Renal Department, University of Uppsala, Uppsala, Sweden); F. Ferrario (MD, Nephropathology Unit, San Gerardo Hospital, Monza, Italy); T. Stellato (MD, Nephropathology Unit, San Gerardo

Hospital, Monza, Italy); J. Egido (MD, Department of Nephrology, Fundacion Jimenez Diaz, Madrid, Spain); C. Martin (MD, Department of Nephrology, Fundacion Jimenez Diaz, Madrid, Spain); J. Floege (MD, Nephrology and Immunology, Medizinische Klinik II, University of Aachen, Aachen, Germany); F. Eitner (MD, Nephrology and Immunology, Medizinische Klinik II, University of Aachen, Aachen, Germany); A. Lupo (MD, Department of Nephrology, University of Verona, Verona, Italy); P. Bernich (MD, Department of Nephrology, University of Verona, Verona, Italy); P. Menè (Department of Nephrology,S. Andrea Hospital, Rome, Italy); M. Morosetti (Nephrology Unit, Grassi Hospital, Ostia, Italy); C. van Kooten, (MD, Department of Nephrology, Leiden University Medical Centre, Leiden, The Netherlands); T. Rabelink (MD, Department of Nephrology, Leiden University Medical Centre, Leiden, The Netherlands); M.E.J. Reinders (MD, Department of Nephrology, Leiden University Medical Centre, Leiden, The Netherlands); J.M. Boria Grinyo (Department of Nephrology, Hospital Bellvitge, Barcelona, Spain); S. Cusinato (MD, Nephrology Unit, Borgomanero Hospital, Borgomanero, Italy); L. Benozzi (MD, Nephrology Unit, Borgomanero Hospital, Borgomanero, Italy); S. Savoldi, (MD, Nephrology Unit, Civile Hospital, Ciriè, Italy); C. Licata (MD, Nephrology Unit, Civile Hospital, Ciriè, Italy); M. Mizerska-Wasiak (MD, Department of Pediatrics, Medical University of Warsaw, Warsaw, Poland); G. Martina (MD, Nephrology Unit, Chivasso Hospital, Chivasso, Italy); A. Messuerotti (MD, Nephrology Unit, Chivasso Hospital, Chivasso, Italy); A. Dal Canton (MD, Nephrology Unit, S. Matteo Hospital, Pavia, Italy); C. Esposito (MD, Nephrology Unit, Maugeri Foundation, Pavia, Italy); C. Migotto (MD, Nephrology Unit, Maugeri Foundation, Pavia, Italy); G. Triolo (MD, Nephrology Unit CTO, Turin, Italy); F. Mariano (MD, Nephrology Unit CTO, Turin, Italy); C. Pozzi (MD, Nephrology Unit, Bassini Hospital, Cinisello Balsamo, Italy); R. Boero (MD, Nephrology Unit, Martini Hospital, Turin, Italy);

VALIGA pathology investigators: S. Bellur (MD, Department of Cellular Pathology, Oxford University Hospitals NHS Foundation Trust, John Radcliffe Hospital, Oxford, United Kingdom); G. Mazzucco (MD, Pathology Department, University of Turin, Turin, Italy); C. Giannakakis (MD, Pathology Department, La Sapienza University, Rome, Italy); E. Honsova (MD, Department of Clinical and Transplant Pathology, Institute for Clinical and Experimental Medicine, Prague, Czech Republic); B. Sundelin (MD Department of Pathology and Cytology, Karolinska University Hospital, Karolinska Institute, Stockholm, Sweden); A.M. Di Palma (Nephrology Unit, Aldo Moro University, Foggia-Bari, Italy); F. Ferrario (MD, Nephropathology Unit, San Gerardo Hospital, Monza, Italy); E. Gutiérrez (MD, Renal, Vascular and Diabetes Research Laboratory, Fundación Instituto de Investigaciones Sanitarias-Fundación Jiménez Díaz, Universidad Autónoma de Madrid, Madrid, Spain); A.M. Asunis (MD, Department of Pathology, Brotzu Hospital, Cagliari, Italy); J. Barratt (MD, The John Walls Renal Unit, Leicester General Hospital, Leicester, United Kingdom); R. Tardanico (MD, Department of Pathology, Spedali Civili Hospital, University of Brescia, Brescia, Italy); A. Perkowska-Ptasinska (MD, Department of Transplantation Medicine, Nephrology and Internal Medicine, Medical University of

Warsaw, Warsaw, Poland); J. Arce Terroba (MD, Pathology Department, Fundació Puigvert, Barcelona, Spain); M. Fortunato (MD, Pathology Department, S. Croce Hospital, Cuneo, Italy); A. Pantzaki (MD, Department of Pathology, Hippokration Hospital, Thessaloniki, Greece); Y. Ozluk (MD, Department of Pathology, Istanbul University, Istanbul Faculty of Medicine, Istanbul, Turkey); E. Steenbergen (MD, Radboud University Medical Center, Department of Pathology, Nijmegen, The Netherlands); M. Soderberg (MD, Department of Pathology, Drug Safety and Metabolism, Huddinge, Sweden); Z. Riispere (MD, Department of Pathology, University of Tartu, Tartu, Estonia); L. Furci (MD, Pathology Department, University of Modena, Italy); D. Orhan (MD, Department of Pediatrics, Division of Rheumatology, Hacettepe University Faculty of Medicine, Ankara, Turkey); D. Kipgen (MD, Pathology Department, Queen Elizabeth University Hospital, Glasgow, United Kingdom); D. Casartelli (Pathology Department, Manzoni Hospital, Lecco, Italy); D. Galesic Ljubanovic (MD, Nephrology Department, University Hospital, Zagreb, Croatia; Zagreb, Croatia); H Gakiopoulou (MD, Department of Pathology, National and Kapodistrian University of Athens, Athens, Greece); E. Bertoni (MD, Nephrology Department, Careggi Hospital, Florence, Italy); P. Cannata Ortiz (MD, Pathology Department, IIS-Fundacion Jimenez Diaz UAM, Madrid, Spain); H. Karkoszka (MD, Nephrology, Endocrinology and Metabolic Diseases, Medical University of Silesia, Katowice, Katowice, Poland); H.J. Groene (MD, Cellular and Molecular Pathology, German Cancer Research Center, Heidelberg, Germany); A. Stoppacciaro (MD, Surgical Pathology Units, Department of Clinical and Molecular Medicine, Ospedale Sant'Andrea, Sapienza University of Rome, Rome, Italy); I. Bajema (MD, Department of Pathology, Leiden University Medical Center, Leiden, The Netherlands); J. Bruijn (MD, Department of Pathology, Leiden University Medical Center, Leiden, The Netherlands); X. Fulladosa Oliveras (MD, Nephrology Unit, Bellvitge University Hospital, Hospitalet de Llobregat, Barcelona, Spain); J. Maldyk (MD, Division of Pathomorphology, Children's Clinical Hospital, Medical University of Warsaw, Warsaw, Poland); and E. Ioachim (MD, Department of Pathology, Medical School, University of Ioannina, Ioannina, Greece); the Oxford derivation and North American validation investigators: Bavbek N (MD, Department of Pathology, Vanderbilt University, Nashville, Tennessee); Cook T (MD, Imperial College, London, England), Troyanov S (MD, Division of Nephrology, Department of Medicine, Hopital du Sacre-Coeur de Montreal, Montreal, Quebec, Canada); Alpers C (MD, Department of Pathology, University of Washington Medical Center, Seattle, Washington), Amore A (MD, Nephrology, Dialysis and Transplantation Unit, Regina Margherita Children's Hospital, University of Turin, Turin, Italy), Barratt J (MD, The John Walls Renal Unit, Leicester General Hospital, Leicester, England); Berthoux F (MD, Department of Nephrology, Dialysis, and Renal Transplantation, Hôpital Nord, CHU de Saint-Etienne, Saint-Etienne, France); Bonsib S (MD, Department of Pathology, LSU Health Sciences Center, Shreveport, Los Angeles); Bruijn J (MD, Department of Pathology, Leiden University Medical Center, Leiden, The Netherlands); D'Agati V (MD, Department of Pathology, Columbia University College of Physicians & Surgeons, New

York, New York); D'Amico G (MD, Fondazione D'Amico per la Ricerca sulle Malattie Renali, Milan, Italy); Emancipator S (MD, Department of Pathology, Case Western Reserve University, Cleveland, Ohio); Emmal F (MD, Division of Nephrology and Dialysis, Department of Nephrology and Urology, Bambino Gesù Children's Hospital and Research Institute, Piazza S Onofrio, Rome, Italy); Ferrario F (MD, Renal Immunopathology Center, San Carlo Borromeo Hospital, Milan, Italy); Fervenza F (MD PhD, Division of Nephrology and Hypertension, Mayo Clinic, Rochester); Florquin S (MD, Department of Pathology, Academic Medical Center, University of Amsterdam, Amsterdam, The Netherlands); Fogo A (MD, Department of Pathology, Vanderbilt University, Nashville, Tennessee); Geddes C (MD, The Renal Unit, Western Infirmary, Glasgow, Scotland); Groene H (MD, Department of Cellular and Molecular Pathology, German Cancer Research Center, Heidelberg, Germany); Haas M (MD, Department of Pathology and Laboratory Medicine, Cedars-Sinai Medical Center, Los Angeles, California); Hill P (MD, St Vincent's Hospital, Melbourne, Australia); Hogg R (MD, Scott and White Medical Center, Temple, Texas (retired)); Hsu S (MD, Division of Nephrology, Hypertension and Renal Transplantation, College of Medicine, University of Florida, Gainesville, Florida); Hunley T (MD, Department of Pathology, Vanderbilt University, Nashville, Tennessee); Hladunewich (MD, Division of Nephrology, Sunnybrook Health Science Center, University of Toronto, Ontario, Canada M); Jennette C (MD, Department of Pathology and Laboratory Medicine, University of North Carolina, Chapel Hill, North Carolina); Joh K (MD, Division of Immunopathology, Clinical Research Center Chiba, East National Hospital, Chiba, Japan); Julian B (MD, Department of Medicine, University of Alabama at Birmingham, Birmingham, Alabama); Kawamura T (MD, Division of Nephrology and Hypertension, Jikei University School of Medicine, Tokyo, Japan); Lai F (MD, The Chinese University of Hong Kong, Hong Kong); Leung C (MD, Department of Medicine, Prince of Wales Hospital, Chinese University of Hong Kong, Hong Kong); Li L (MD, Research Institute of Nephrology, Jinling Hospital, Nanjing University School of Medicine, Nanjing, China); Li P (MD, Department of Medicine, Prince of Wales Hospital, Chinese University of Hong Kong, Hong Kong); Liu Z (MD, Research Institute of Nephrology, Jinling Hospital, Nanjing University School of Medicine, Nanjing, China); Massat A (MD, Division of Nephrology and Hypertension, Mayo Clinic, Rochester, Minnesota); Mackinnon B (MD, The Renal Unit, Western Infirmary, Glasgow, Scotland); Mezzano S (MD, Departamento de Nefrología, Escuela de Medicina, Universidad 5 Austral, Valdivia, Chile); Schena F (MD, Renal, Dialysis and Transplant Unit, Policlinico, Bari, Italy); Tomino Y (MD, Division of Nephrology, Department of Internal Medicine, Juntendo University School of Medicine, Tokyo, Japan); Walker P (MD, Nephropathology Associates, Little Rock, Arkansas); Wang H (MD, Renal Division of Peking University First Hospital, Peking University Institute of Nephrology, Beijing, China (deceased)); Weening J (MD, Erasmus Medical Center, Rotterdam, The Netherlands); and Yoshikawa N (MD, Department of Pediatrics, Wakayama Medical University, Wakayama City, Japan);

International investigators: Cai-Hong Zeng (MD, Nanjing University School of Medicine, Nanjing, China); Sufang Shi (MD, Peking University Institute of Nephrology, Beijing, China); C. Nogi (MD, Juntendo University, Faculty of Medicine, Tokyo, Japan); H. Suzuki (MD, Juntendo University, Faculty of Medicine, Tokyo, Japan); K. Koike (MD, Division of Nephrology and Hypertension, Department of Internal Medicine, Jikei University School of Medicine, Tokyo, Japan); K. Hirano (MD, Division of Nephrology and Hypertension, Department of Internal Medicine, Jikei University School of Medicine, Tokyo, Japan); T. Kawamura (MD, Division of Nephrology and Hypertension, Department of Internal Medicine, Jikei University School of Medicine, Tokyo, Japan); T. Yokoo (MD, Division of Nephrology and Hypertension, Department of Internal Medicine, Jikei University School of Medicine, Tokyo, Japan); M. Hanai (MD, Division of Nephrology, Department of Medicine, Kurume University School of Medicine, Fukuoka, Japan); K. Fukami (MD, Division of Nephrology, Department of Medicine, Kurume University School of Medicine, Fukuoka, Japan); K. Takahashi (MD, Department of Nephrology, Fujita Health University School of Medicine, Aichi, Japan); Y. Yuzawa (MD, Department of Nephrology, Fujita Health University School of Medicine, Aichi, Japan); M. Niwa (MD, Department of Nephrology, Nagoya University Graduate School of Medicine, Aichi, Japan); Y. Yasuda (MD, Department of Nephrology, Nagoya University Graduate School of Medicine, Aichi, Japan); S. Maruyama (MD, Department of Nephrology, Nagoya University Graduate School of Medicine, Aichi, Japan); D. Ichikawa (MD, Division of Nephrology and Hypertension, Department of Internal Medicine, St Marianna University School of Medicine, Kanagawa, Japan); T. Suzuki (MD, Division of Nephrology and Hypertension, Department of Internal Medicine, St Marianna University School of Medicine, Kanagawa, Japan); S. Shirai (MD, Division of Nephrology and Hypertension, Department of Internal Medicine, St Marianna University School of Medicine, Kanagawa, Japan); A. Fukuda (MD, First Department of Internal Medicine, Faculty of Medicine, University of Miyazaki, Miyazaki, Japan); S. Fujimoto (MD, Department of Hemovascular Medicine and Artificial Organs, Faculty of Medicine, University of Miyazaki, Miyazaki, Japan); H. Trimarchi (MD, Division of Nephrology, Hospital Britanico, Buenos Aires, Argentina).

**The Cure Glomerulonephropathy (CureGN) investigators are as follows:**

CureGN collaborators: The CureGN Consortium members listed below, from within the four Participating Clinical Center networks and Data Coordinating Center, are acknowledged by the authors as Collaborators.

**CureGN PCC Principal Investigators; *CureGN Site Principal Investigators; +CureGN Pathologists, #CureGN Lead Coordinators.

CureGN Participating Clinical Centers (PCC) through Columbia University:

*Columbia University, New York, NY, US*: Gerald Appel, Revekka Babayev, Ibrahim Batal +, Andrew Bomback**, Pietro Canetta, Brenda Chan, Vivette Denise D'Agati +, Samitri Dogra, Hilda Fernandez, Gabriele Gaggero+, Ali Gharavi**, William Hines, , Krzysztof Kiryluk**, Satoru Kudose +, Fangming Lin, Victoria Kolupaeva#, Maddalena Marasa, Glen Markowitz +, Mariela Navarro-Torres, Hila Milo Rasouly, Sumit Mohan, Nicola Mongera,

Jordan Nestor, Jai Radhakrishnan, Maya Rao, Maya Sabatello, Simone Sanna-Cherchi, Dominick Santoriello+, Miroslav Sekulic +, , Michael Barry Stokes+, Natalie Uy, Natalie Vena, Benjamin Wooden

*University of Warsaw, Warszawa, Poland:* Bartosz Foroncewicz, Natalia Wiewiórska-Krata, Barbara Moszczuk, Krzysztof Mucha*, Agnieszka Perkowska-Ptasińska, Elżbieta Ryszkowska

*IRCCS Giannina Gaslini, Genoa, Italy*: Francesca Lugani, Valerio Vellone+

<u>CureGN Participating Clinical Centers (PCC) through the Pediatric Nephrology Research Consortium:</u>

*Children's Hospital of New Orleans/ LSU Health, New Orleans, LA, USA*: Diego Aviles*

*Children's Mercy Hospital, Kansas City, MO, USA*: Tarak Srivastava*, Alexander Katz+

*Children's National Medical Center, Washington DC, USA*: Sun-Young Ahn*

*Cincinnati Children's Hospital Cincinnati, OH, USA*: Prasad Devarajan, Elif Erkan*, Hillarey Stone

*Connecticut Children's Medical Center, Hartford, CT, USA*: Sherene Mason*

*East Carolina University Brody School of Medicine, Greenville, NC, USA*: Liliana Gomez-Mendez*

*Emory University, Atlanta, GA, USA*: Larry Greenbaum**, Chia-shi Wang, Hong (Julie) Yin+

*Helen DeVos Children's Hospital, Grand Rapids, MI, USA*: Goebel Jens*

*Levine Children's Hospital/Atrium Health, Charlotte, NC, USA*: Donald Weaver*

*Lurie Children's Hospital, Chicago IL, USA*: Jill Krissberg*, Jerome Lane

*Medical College of Wisconsin, Milwaukee, WI, USA*: Cindy Pan, Ellen Cody*

*Nationwide Children's Hospital, Columbus, OH, USA*: Samantha Martinek-Bundt#, Dawson Carmean#, Mary Dreher#, Mahmoud Kallash*, John Mahan**, Samantha Sharpe#, William Smoyer**, Laura Biederman+

*Oregon Health and Science University, Portland, OR, USA*: Amira Al-Uzri*, Sandra Iragorri

*Riley Children's Hospital, Indianapolis, IN, USA*: Myda Khalid**

*Cardinal Glennon Children's Medical Center/ St. Louis University, St. Louis, MO, USA*: Craig Belsha*

*Texas Children's Hospital, Houston, TX, USA*: Elizabeth Onugha*, Michael Braun, AC Gomez

*Texas Tech Health Sciences Center, Amarillo, TX, USA*: Tetyana Vasylyeva*

*Children's of Alabama, University of Alabama, Birmingham, AL, USA*: Daniel Feig*

*University of Colorado Children's Hospital, Colorado, Aurora, CO, USA*: Melisha Hannah*

*University of Kentucky, Lexington, KY, USA*: Aftab Chishti*

*University of Louisville, Louisville, KY, USA*: Jon Klein**

*Holtz Medical Center, University of Miami, Miami, FL, USA*: Chryso Katsoufis, Wacharee Seeherunvong*

*University of Minnesota Children's Hospital, Minneapolis, MN, USA*: Michelle Rheault**

*University of New Mexico Health Sciences Center, Albuquerque, NM, USA*: Craig Wong*

*University of Oklahoma Health Sciences Center, Oklahoma City, OK, USA*: Qassim Abid*
*University of Virginia, Charlottesville, VA, USA*: John Barcia*, Agnes Swiatecka-Urban
*University of Wisconsin, Madison, WI, USA*: Sharon Bartosh*
*Washington University in St. Louis, St. Louis, MO, USA*: Brian Stotter*, Joseph Gaut [+]
CureGN Participating Clinical Centers (PCC) through the University of North Carolina:
*Hôpital Maisonneuve-Rosemont, Montreal, Canada*: Louis-Philippe Laurin*, Virginie Royal[+], Mathieu Latour[+], Natlie (Natacha) Patey[+]
*Medical University of South Carolina, Charleston, SC, USA*: Anand Achanti, Milos Budisavljevic*, Vishwajeeth Pasham[+]
*Northwestern University, Chicago, IL, USA*: Cybele Ghossein, Yonatan Peleg*
*Ohio State University, Columbus, OH, USA*: Salem Almaani*, Isabelle Ayoub, Samir Parikh, Brad Rovin, Anjali Satoskar[+]
*University of Chicago, Chicago, IL, USA*: Anthony Chang[+]
*University of Alabama at Birmingham, Birmingham, AL, USA*: Huma Fatima[+], Jan Novak, Matthew Renfrow, Dana Rizk*
*University of North Carolina Kidney Center, Chapel Hill, NC, USA*: Dhruti Chen, Vimal Derebail**, Ronald Falk**, Keisha Gibson, Dorey Glenn, Susan Hogan, Koyal Jain, J. Charles Jennette[+], Vanessa Moreno[+], Amy Mottl, Caroline Poulton[#], Monica Reynolds, Manish Kanti Saha, Nicole E. Wyatt
*Vanderbilt University, Nashville, TN, USA*: Agnes Fogo[+], Neil Sanghani*
*Virginia Commonwealth University, Richmond, VA, USA*: Jason Kidd*, Selvaraj Muthusamy[+]
CureGN Participating Clinical Centers (PCC) through the University of Pennsylvania:
*Children's Hospital of Philadelphia, Philadelphia, PA, USA*: Rebecca Scobell*, Michelle Denburg, Amy Kogon, Kevin Meyers, Madhura Pradhan
*Cleveland Clinic, Cleveland, OH, CA*: Raed Bou Matar*, John O'Toole, John Sedor
Cohen Children's Medical Center, New Hyde Park, NY, USA: Christine Sethna**, Suzanne Vento[#]
*Johns Hopkins University, Baltimore, MD, USA*: Mohamed Atta, Serena Bagnasco[+], Alicia Neu, John Sperati*
*Lundquist Institute at Harbor-UCLA Medical Center, Torrance, CA, USA*: Sharon Adler*, Tiane Dai, Ram Dukkipati
*Montefiore Medical Center, The Bronx, New York, NY, USA*: Frederick Kaskel, Kaye Brathwaite, Kimberly Reidy*
*New York University, New York, NY, USA*: Laura Malaga-Dieguez*
*Spokane Providence Medical Center, Spokane, WA, USA*: Katherine Tuttle*
*Stanford University, Palo Alto, CA, USA*: Richard Lafayette*, Kamal Fahmeedah, Elizabeth Talley
*Sunnybrook Health Sciences Centre, Toronto, Canada*: Michelle Hladunewich*
*The Hospital for Sick Children, Toronto, Canada*: Rulan Parekh*

*University Health Network, Toronto, Canada*: Carmen Avila-Casado[+], Daniel Cattran*, Reich Heather, Meherzad Kutky
*University of Miami, Miami, FL, USA*: Yelena Drexler*, Alessia Fornoni
*University of Michigan, Ann Arbor, MI, USA*: Jeffrey Hodgin[+], Andrea Oliverio*
*University of Pennsylvania, Philadelphia, PA, USA*: Jon Hogan, Lawrence Holzman**, Matthew Palmer [+], Gaia Coppock
*University of Pittsburgh School of Medicine, Pittsburgh, PA, USA*: Michael Mortiz, Juhi Kumar*
*University of Washington, Seattle, WA, USA*: Charles Alpers[+], J. Ashley Jefferson*
*UT Southwestern, Dallas, TX, USA*: Kamal Sambandam, Bethany Roehm*
Data Coordinating Center (DCC):
*Cedar Sinai Medical Center, Los Angeles, CA, USA*: Cynthia Nast[+], Jean Hou[+]
*Duke University, Durham, NC, USA*: Laura Barisoni
*Cleveland Clinic, Cleveland, OH, USA:* Crystal Gadegbeku**
*Northwestern University, Chicago, IL, USA:* Abigail Smith**
*University of Michigan, Ann Arbor, MI, USA*: Brenda Gillespie, Bruce Robinson, Matthias Kretzler, Zubin Modi, Laura Mariani**
Steering Committee Chair: Lisa M. Guay-Woodford, Children's Hospital of Pennsylvania, Philadelphia, PA, USA.

**The National Unified Renal Translational Research Enterprise (NURTuRE) academic steering group members are as follows:**
NURTuRE academic steering group: M.W. Taal (Centre for Kidney Research and Innovation, University of Nottingham, Derby, United Kingdom and University Hospitals of Derby and Burton NHS Foundation Trust, Derby, United Kingdom), P. Cockwell (Department of Renal Medicine, Queen Elizabeth Hospital, University Hospitals of Birmingham, Birmingham, UK and Institute of Inflammation and Ageing, University of Birmingham, UK), S.D.S. Fraser (School of Primary Care, Population Sciences and Medical Education, Faculty of Medicine, University of Southampton, Southampton, UK), P.A. Kalra (Donal O'Donoghue Renal Research Centre, Salford Royal Hospital, Northern Care Alliance NHS Foundation Trust, Salford, UK and University of Manchester, Faculty of Biology Medicine and Health, Division of Cardiovascular Sciences, Oxford Rd, Manchester, UK), M. Saleem (Bristol Renal and Children's Renal Unit, Bristol Medical School, University of Bristol, Bristol, UK), D.C. Wheeler (Department of Renal Medicine, University College London, London, UK)

**The AI4IgAN study members are as follows:**
AI4IgAN investigators: R. Coppo for the VALIGA study (MD, Regina Margherita Childrens University Hospital, Turin, Italy); S. Barbour (MD, Division of Nephrology, Department of Medicine, University of British Columbia, Vancouver, Canada); J. Barratt (MD, The John Walls Renal Unit, Leicester General Hospital, Leicester, United Kingdom); I.S.D. Roberts (MD, Department of Cellular Pathology, Oxford University Hospitals NHS Foundation

Trust, John Radcliffe Hospital, Oxford, United Kingdom); J. Floege (MD, Nephrology, Immunology and Cardiology, Medizinische Klinik I & II, University of Aachen, Aachen, Germany); V. Tesar (MD, PhD, Department of Nephrology, 1st Faculty of Medicine and General University Hospital, Charles University, Prague, Czech Republic); R.D. Bülow (MD, Institute of Pathology, University of Aachen, Aachen, Germany); H. Zhang (MD, PhD, Renal Division, Peking University First Hospital, Peking, China); M.G. Wong (MD, PhD, Department of Renal Medicine, Royal North Shore Hospital, Sydney, Australia); L. Barisoni (MD, Department of Medicine, Division of Nephrology, Mayo Clinic, Rochester, Minnesota, USA & Department of Pathology, Division of AI and Computational Pathology, Duke University, Durham, North Carolina, USA); M. Haas (MD, Department of Pathology & Laboratory Medicine, Cedars-Sinai Medical Center, Los Angeles, California, USA); M. Yanagita (MD, PhD, Department of Nephrology, Kyoto University, Kyoto, Japan); K. Kaneko (MD, Department of Nephrology, Kyoto University, Kyoto, Japan); T. Koshida (MD, Department of Nephrology, Faculty of Medicine, Juntendo University, Tokyo, Japan); S. Lundberg (MD, PhD, Department of Nephrology, Karolinska Institutet/Danderyd University Hospital, Stockholm, Sweden).

### Supplementary Methods

#### Cohort refinement

All eight cohorts included in this study were collected retrospectively, independently and autonomously for research purposes. All samples across cohorts were subject to the same defined inclusion criteria. We included patients ≥ 14 years of age with biopsy-proven IgA nephropathy (IgAN) without any reported secondary causes (e.g., liver disease) or IgA vasculitis, no kidney failure at time of biopsy, no missing estimated glomerular filtration rate (eGFR) at time of biopsy and at least one month of clinical follow-up.

To maintain consistency across cohorts and minimize treatment-related confounding, patients who were enrolled in the respective study over six months after kidney biopsy, received documented corticosteroid treatment prior to kidney biopsy or only after one year of follow-up, did receive other immunosuppressive agents (e.g., cyclophosphamide), targeted-release corticosteroid formulations (e.g., Nefecon) or dual endothelin and angiotensin II receptor antagonists (DERAs, e.g., Sparsentan) were excluded. Cases without sufficient number of glomeruli according to the Oxford classification for IgA nephropathy[1] or with broken/damaged glass slides were also excluded. As multiple studies have shown a considerably increased rate in serious adverse events (SAEs) in patients with decreased kidney function ≤30ml/min/1.73m$^2$[2,3] these patients were also excluded.

The European Validation Study of the Oxford Classification of IgA Nephropathy (VALIGA) study[4] is a multi-center cohort across 55 participating centers in Europe including a secondary analysis of corticosteroid therapy[5]. In total, 507 patients met the above-mentioned criteria and were considered for further analysis.

The Kyoto cohort (n= 92) is a single-center cohort from the Kyoto University Hospital in Japan with kidney biopsies taken between 2013 and 2020.

The National Unified Renal Translational Research Enterprise chronic kidney disease (NURTuRE-CKD) cohort[6] (n = 20) is a national multi-centric prospective cohort of 2,996 participants from 16 participating hospitals in the United Kingdom, biopsy samples were collected between 2017 to 2019.

The Leicester cohort (n = 93) is a single-center cohort from the University Hospital of Leicester National Health Service Trust in the United Kingdom with kidney biopsies performed from 2010 to 2023.

The Aachen cohort (n = 27) is a single-center cohort from the University Hospital RWTH Aachen in Germany with kidney biopsies from 2018 to 2025.

The Diyarbakir cohort (n = 106) is a single-center cohort from the Dicle University Hospital in Turkey. Kidney biopsies were collected from 2015 to 2024.

The Rochester cohort (n = 42) is a single-center cohort of local patients at the Mayo Clinic in Rochester (United States of America) with kidney biopsies performed between 1996 and 2015[7].

The Cure Glomerulonephropathy (CureGN) cohort (n = 135) is a multi-centric observational study of glomerular disease including IgAN[8]. Patients were recruited from 57 study sites in the United States of America, Canada, Italy and Poland.

In total, 1,022 eligible patients were identified and divided into derivation and validation cohorts. To ensure a similar baseline hazard, covariates and treatment assignment percentage, the VALIGA cohort was randomly split by clinical center into derivation and validation subsets[9]. In summary, 464 patients from the VALIGA, Kyoto and NURTuRE-CKD cohorts formed the derivation cohort while 558 patients from the VALIGA, Leicester, Aachen, Diyarbakir, Rochester and CureGN cohorts formed the validation cohort.

#### Definitions for predictors

Proteinuria, mean arterial blood pressure (MAP) and eGFR at kidney biopsy were reported as the closest values to the time of biopsy within a 180-day period. For adults, eGFR was calculated from serum creatinine using the 2021 CKD-EPI equation[10], while for children (<18 years of age), the Schwartz formula[11] was applied (as done previously in VALIGA[5]). Proteinuria was reported as grams per day and taken from 24-hour urine collections. Whenever these were not available, the daily urinary protein excretion was estimated from spot urine protein or albumin to creatinine ratios by using previously validated regression equations[12,13]. Reported age corresponds to age at time of biopsy. Race and ethnicity were collected based on reports from individual databases in accordance with local guidelines (e.g., National Health Service Ethnicity Codes[14]) and summarized into the categories ‘White’, ‘Black’, ‘South Asian’, ‘Chinese’, ‘Japanese’ and ‘Other’ (e.g., Pacific

Islanders, Hispanic)[15]. Although these categories might not fully represent the granularities between patients and cannot account for social or other demographic factors, they were designed to capture potential treatment effect modifiers, as different progression rates and responses to immunosuppressive treatments have been reported in these populations[16].

All included biopsy cases were scored locally in accordance with the Oxford classification for IgAN[1,17], missing cases were rescored centrally blinded to clinical and other pathological data. The MEST-C score components were defined as follows: M0/M1 as mesangial hypercellularity (≥ 4 mesangial cells per mesangial area) with < or ≥ 50% of glomeruli affected, E1/E0 as the presence or absence of endocapillary hypercellularity in any glomerulus, S1/S0 as the presence or absence of segmental sclerosis and/or tuft adhesions in any glomerulus and T0/T1/T2 as the degree of tubular atrophy and/or interstitial fibrosis of the cortical area (< 25%, 25-50%, > 50%). Crescents were only considered as the absence or presence cellular of fibrocellular crescents (C0/C1) due to the low incidence of ≥ 25% glomeruli with crescents (C2) in the derivation cohort.

The primary composite outcome was defined as progression of IgAN. This included a sustained ≥ 50% decline in eGFR and/or development of kidney failure, including persistent eGFR below 15ml/min/1.73m², initiation of dialysis or kidney transplantation within five years of follow-up after time of kidney biopsy as this corresponds to the median follow-up duration for both cohorts[18,19]. Outcomes were analyzed using pseudo-values for the restricted mean survival time (RMST) as the area under the survival curve[20]. This approach allows for a more flexible and robust effect modelling for right-censored survival analysis[21].

For all patients' information on corticosteroid and renin-angiotensin system blocker (RASi) treatment during follow-up were collected, although reporting of dosage and formulations was inconsistent.

Missingness in baseline predictors was limited and observed only for proteinuria and MAP (Supplementary Table 11). To address this, we performed multivariate imputation via chained equations (MICE)[22–24] using predictive mean matching for continuous variables with 20 multiple imputations and 10 iterations.

#### Computation of pathomics features

Kidney biopsies from all included patients were digitized and analyzed using our previously published framework for pathomics in Nephropathology[25]. For better segmentation accuracy across all cohorts, we refined our previous approach to using a MedNeXt-based architecture[26] with subsequent computation of quantitative morphometric features on instance-level. All features were then aggregated at biopsy-level using the median of the distribution[27]. Definitions for the five pathomics predictors are provided in Supplementary Table 9.

#### Model architecture

To estimate individualized treatment effects (ITEs) from observational data, we implemented the X-learner, a meta-learning algorithm designed for causal inference. The X-learner proceeds in three main steps. First, two outcome models are trained separately: one for the untreated group to estimate

$$\mu_0(X)=[Y|T=0, X],$$

and one for the treated group to estimate

$$\mu_1(X)=[Y|T=1, X],$$

where X denotes covariates, Y the outcome and T the treatment. These models predict potential outcomes for all individuals under both treatment conditions. In the second step, individual-level pseudo-treatment effects are imputed: for treated individuals, the observed outcome is compared to their predicted control outcome

$$\tau(X, T=1)=YT = 1-\mu_0(X),$$

and for controls, the predicted treated outcome is compared to the observed control outcome

$$\tau(X, T=0)=\mu 1(X)\text{-}YT=0.$$

These differences serve as pseudo-outcomes that are then modeled separately in each group as functions of patient covariates. In the final step, the two resulting treatment effect models are combined to form a final estimate of $\tau(X)$, using weights derived from a propensity score model that reflects the probability of being treated given covariates. This weighted averaging enhances stability, particularly in settings where the proportion of treated and untreated individuals is imbalanced.

In our implementation, gradient-boosted trees (XGBoost) were used as base learners for both outcome and treatment effect modeling due to their robustness to nonlinearities and high-dimensional data. The framework was implemented using the *rlearner* package in R[28,29].

#### Model assessment

To assess whether our model reliably captured treatment heterogeneity, we evaluated multiple performance metrics: the Qini coefficient, the rank-weighted average treatment effect (RATE) and the concordance statistic for benefit (C-for-benefit). For each metric, 95% confidence intervals were estimated via bootstrap resampling.

*Qini*. The Qini curve quantifies the added benefit of targeting corticosteroid therapy based on predicted ITEs. Patients are ranked by decreasing ITE (i.e., from highest to lowest benefit), partitioned into equal-sized groups, and cumulative incremental benefit is computed within each group relative to a baseline strategy of random treatment allocation. To summarize the model's discriminative ability in relation to the baseline we compute the Qini coefficient (area between the Qini curve and reference line[30–32]).

*Rank-weighted average treatment benefit (RATE)*. The RATE curve evaluates model ranking by estimating the average treatment effect (ATE) among fractions of the population ordered by predicted ITE. Patients are binned by rank, group-level ATEs are computed, and cumulative averages across bins are plotted. The overall RATE corresponds to the area under the RATE curve, reflecting the degree to which the model successfully ranks patients by true treatment benefit[33,34].

*Concordance statistic for benefit (C-for-benefit)*. The C-for-benefit quantifies the model's ability to discriminate between patients with higher versus lower observed treatment effects[35,36]. It estimates the probability that, for randomly selected patients, the individual with the higher predicted ITE also had a larger observed benefit. C-for-benefit values above 0.5 indicate that the model can distinguish between these patients better than random chance, with higher values indicating better discrimination. Due to the unbalanced treatment assignment patients were matched with two controls per treated (2:1 matching). We further assessed overall model calibration in the validation cohort. Calibration refers to the agreement between the observed and predicted treatment benefit with optimal calibration referring to both being equal.

#### Sensitivity and robustness analyses

After completing model derivation, we conducted multiple sensitivity and robustness analyses to validate our findings. To account for any potential cohort-bias we introduced an interaction term to assess whether the cohort modified the observed or predicted treatment effect in all patients. Additionally, the corticosteroid treatment duration and race and ethnicity as potential treatment effect modifiers were similarly assessed by linear regression and likelihood ratio testing.

Furthermore, we added four robustness checks. (1) *Random predictor:* a randomly generated binary predictor was added to the derivation matrix. (2) *Random replace:* proteinuria was replaced by a randomly generated continuous predictor with a similar range of distribution. (3) *Random treatment:* the binary corticosteroid treatment assignment was changed to a random treatment assignment with similar treatment percentage. (4) *Random outcome:* the RMST was replaced by a random outcome with similar range to the observed RMST in the derivation cohort. For each check, the model was refit and model robustness was assessed by comparing the predicted ATE with 95% confidence intervals by bootstrapping in the derivation cohort.

#### Scoring of tubulointerstitial inflammation

To further assess the importance of combining tubulointerstitial pathomics features, 60 cases with increased tubular distance were scored by a trained nephropathologist for interstitial inflammation (*i*) and tubulitis (*t*) in accordance with the Banff classification for kidney transplant pathology[37]. Interstitial inflammation was defined as i0: no inflammation or in less than 10% of unscarred cortical parenchyma, i1: inflammation in 10 to 25% of unscarred cortical parenchyma, i2: inflammation in 26 to 50% of unscarred cortical parenchyma and i3: inflammation in more than 50% of unscarred cortical parenchyma. Tubulitis was defined as t0: no mononuclear cells in tubules or single focus of tubulitis only, t1: two or more foci with one to four mononuclear cells/tubular cross section (or ten tubular cells) in the most affected tubule, t2: two or more foci of tubulitis, at least one of those foci with five to ten mononuclear cells/tubular cross section (or ten tubular cells) in the most affected tubule and t3: two or more foci of tubulitis, at least one of those foci with over ten mononuclear cells/tubular cross section in the most affected tubule, or the presence of ≥2 areas of tubular basement membrane destruction accompanied by i2/i3 inflammation and t2 elsewhere. Each half of the cases represented cases with high or low predicted corticosteroid treatment effect, and all cases were scored in blinded fashion without any knowledge of predicted treatment benefit, associated clinical or pathomics data.

### Supplementary Tables and Figures

**Supplementary Table 1.** Additional baseline characteristics of the derivation and validation cohort. Continuous variables are reported as median (interquartile range) while categorical variables are reported as absolute (n) and relative (%) frequencies.

| | **Derivation** | **Validation** |
|---|---|---|
| n | 464 | 558 |
| Systolic blood pressure [mmHg] | 130.0 (20.0) | 122.5 (27.0) |
| Diastolic blood pressure [mmHg] | 80.0 (19.0) | 78.0 (16.0) |
| BMI [kg/m²] | 24.3 (5.6) | 25.7 (6.6) |
| Race and ethnicity | White = 348 (75.8%)<br>Black = 1 (0.2%)<br>South Asian = 13 (2.8%)<br>Chinese = 0 (0.0%)<br>Japanese = 92 (20.1%)<br>Other = 5 (1.1%) | White = 446 (91.0%)<br>Black = 5 (1.0%)<br>South Asian = 27 (5.5%)<br>Chinese = 4 (0.8%)<br>Japanese = 0 (0.0%)<br>Other = 8 (1.7%) |

**Supplementary Table 2.** Baseline patient characteristics in the individual cohorts. Continuous variables are reported as median (interquartile range) while categorical variables are reported as absolute (n) and relative (%) frequencies.
Abbreviations: m, male; f, female; eGFR, estimated glomerular filtration rate; MAP, mean arterial blood pressure; M, mesangial hypercellularity; E, endocapillary hypercellularity; S, segmental glomerulosclerosis; T, tubular atrophy and interstitial fibrosis; C, crescents; RASi, renin-angiotensin-system inhibitor; CS, corticosteroid.

| | **VALIGA** | **Kyoto** | **NURTuRE-CKD** | **Leicester** | **Aachen** | **Diyarbakir** | **Rochester** | **CureGN** |
|---|---|---|---|---|---|---|---|---|
| n | 507 | 92 | 20 | 93 | 27 | 106 | 42 | 135 |
| Follow-up [years] | 5.1 (6.3) | 2.5 (2.9) | 4.1 (1.4) | 7.7 (5.6) | 1.3 (3.3) | 1.7 (2.6) | 13.9 (9.3) | 6.3 (2.8) |
| Age [years] | 34.5 (20.8) | 40.2 (22.4) | 50.0 (16.3) | 39.6 (19.8) | 34.0 (25.5) | 33.0 (16.8) | 42.0 (20.5) | 38.3 (19.7) |
| Sex [m\|f] | m: 381 (75.1%) \| f: 126 (24.9%) | m: 39 (42.4%) \| f: 53 (57.6%) | m: 13 (65.0%) \| f: 7 (35.0%) | m: 60 (64.5%) \| f: 33 (35.5%) | m: 19 (70.4%) \| f: 8 (29.6%) | m: 54 (50.9%) \| f: 52 (49.1%) | m: 29 (69.0%) \| f: 13 (31.0%) | m: 85 (63.0%) \| f: 50 (37.0%) |
| eGFR [ml/min/1.73m²] | 81.4 (46.5) | 97.5 (48.8) | 46.6 (15.3) | 71.5 (51.5) | 71.0 (33.6) | 86.8 (66.5) | 73.1 (33.7) | 66.0 (53.4) |
| Proteinuria [g/24h] | 1.0 (1.5) | 0.6 (1.1) | 1.2 (1.3) | 0.8 (1.2) | 1.0 (0.9) | 2.0 (3.3) | 1.3 (1.6) | 1.2 (1.6) |
| MAP [mmHg] | 96.7 (16.7) | 90.2 (15.1) | 100.0 (19.1) | 95.3 (20.0) | 92.0 (16.0) | 88.5 (13.3) | 99.5 (22.5) | 94.7 (15.0) |
| M [0\|1] | 0: 369 (72.8%) \| 1: 138 (27.2%) | 0: 50 (54.3%) \| 1: 42 (45.7%) | 0: 7 (35.0%) \| 1: 13 (65.0%) | 0: 32 (34.4%) \| 1: 61 (65.6%) | 0: 18 (66.7%) \| 1: 9 (33.3%) | 0: 72 (67.9%) \| 1: 34 (32.1%) | 0: 3 (7.1%) \| 1: 39 (92.9%) | 0: 11 (8.1%) \| 1: 124 (91.9%) |
| E [0\|1] | 0: 454 (89.5%) \| 1: 53 (10.5%) | 0: 68 (73.9%) \| 1: 24 (26.1%) | 0: 9 (45.0%) \| 1: 11 (55.0%) | 0: 68 (73.1%) \| 1: 25 (26.9%) | 0: 18 (66.7%) \| 1: 9 (33.3%) | 0: 87 (82.1%) \| 1: 19 (17.9%) | 0: 38 (90.5%) \| 1: 4 (9.5%) | 0: 71 (52.6%) \| 1: 64 (47.4%) |
| S [0\|1] | 0: 138 (27.2%) \| 1: 369 (72.8%) | 0: 6 (6.5%) \| 1: 86 (93.5%) | 0: 5 (25.0%) \| 1: 15 (75.0%) | 0: 21 (22.6%) \| 1: 72 (77.4%) | 0: 6 (22.2%) \| 1: 21 (77.8%) | 0: 91 (85.8%) \| 1: 15 (14.2%) | 0: 17 (40.5%) \| 1: 25 (59.5%) | 0: 4 (3.0%) \| 1: 131 (97.0%) |
| T [0\|1\|2] | 0: 416 (82.0%) \| 1: 84 (16.6%) \| 2: 7 (1.4%) | 0: 67 (72.8%) \| 1: 24 (26.1%) \| 2: 1 (1.1%) | 0: 15 (75.0%) \| 1: 4 (20.0%) \| 2: 1 (5.0%) | 0: 84 (90.3%) \| 1: 8 (8.6%) \| 2: 1 (1.1%) | 0: 24 (88.9%) \| 1: 2 (7.4%) \| 2: 1 (3.7%) | 0: 37 (34.9%) \| 1: 64 (60.4%) \| 2: 5 (4.7%) | 0: 34 (80.9%) \| 1: 7 (16.7%) \| 2: 1 (2.4%) | 0: 75 (55.5%) \| 1: 58 (43.0%) \| 2: 1 (1.5%) |

| | | | | | | | | |
|---|---|---|---|---|---|---|---|---|
| C [0\|1] | 0: 456 (89.9%) \| 1: 51 (10.1%) | 0: 68 (73.9%) \| 1: 24 (26.1%) | 0: 13 (65.0%) \| 1: 7 (35.0%) | 0: 65 (69.9%) \| 1: 28 (30.1%) | 0: 23 (85.2%) \| 1: 4 (14.8%) | 0: 100 (94.3%) \| 1: 6 (5.7%) | 0: 35 (83.3%) \| 1: 7 (16.7%) | 0: 82 (60.7%) \| 1: 53 (39.3%) |

**Supplementary Table 3.** Restricted mean progression-free survival (RMST) based on treatment policy.
Abbreviations: R, recommended; T, treated; CS, corticosteroid.

| Cohort | Group | RMST | Difference in RMST |
|---|---|---|---|
| Derivation | R = no CS \| T = no CS | 4.97 [95% CI 4.93-5.01] | 0.17 [95% CI 0.01-0.33], $p = 0.22$ |
| | R = no CS \| T = CS | 4.8 [95% CI 4.64-4.95] | |
| | R = CS \| T = CS | 4.84 [95% CI 4.64-5.04] | 0.56 [95% CI 0.23-0.89], $p < 0.01$ |
| | R = CS \| T = no CS | 4.28 [95% CI 4.02-4.54] | |
| | R ≠ T | 4.51 [95% CI 4.35-4.68] | 0.43 [95% CI 0.26-0.6], $p < 0.01$ |
| | R = T | 4.94 [95% CI 4.89-4.99] | |
| Validation | R = no CS \| T = no CS | 4.81 [95% CI 4.71-4.9] | 0.01 [95% CI -0.2-0.23], $p = 1.00$ |
| | R = no CS \| T = CS | 4.79 [95% CI 4.6-4.99] | |
| | R = CS \| T = CS | 4.88 [95% CI 4.71-5.05] | 0.4 [95% CI 0.13-0.67], $p < 0.05$ |
| | R = CS \| T = no CS | 4.48 [95% CI 4.27-4.69] | |
| | R ≠ T | 4.58 [95% CI 4.42-4.73] | 0.24 [95% CI 0.06-0.42], $p < 0.01$ |
| | R = T | 4.82 [95% CI 4.73-4.9] | |

**Supplementary Table 4.** Derivation cohort: characteristics of partitioned subgroups. Continuous variables are reported as median (interquartile range) while categorical variables are reported as absolute (n) and relative (%) frequencies. The primary outcome was defined as a composite of ≥ 50% eGFR reduction and kidney failure within five years of kidney biopsy.
Abbreviations: R, recommended; T, treated; CS, corticosteroid; m, male; f, female; eGFR, estimated glomerular filtration rate; MAP, mean arterial blood pressure; M, mesangial hypercellularity; E, endocapillary hypercellularity; S, segmental glomerulosclerosis; T, tubular atrophy and interstitial fibrosis; C, crescents; RASi, renin-angiotensin-system inhibitor.

| | **R = no CS \| T = no CS** | **R = no CS \| T = CS** | **R = CS \| T = CS** | **R = CS \| T = no CS** | **R ≠ T** | **R = T** |
|---|---|---|---|---|---|---|
| n | 202 | 107 | 52 | 103 | 210 | 254 |
| Follow-up [years] | 6.0 (7.5) | 3.1 (3.7) | 3.8 (2.7) | 4.6 (6.9) | 4.0 (4.3) | 5.2 (6.7) |
| Age [years] | 33.1 (23.0) | 36.7 (18.5) | 39.8 (21.2) | 37.0 (20.2) | 36.8 (20.5) | 34.1 (22.3) |
| Sex [m\|f] | m: 136 (67.3%) \| f: 66 (32.7%) | m: 63 (58.9%) \| f: 44 (41.1%) | m: 33 (63.5%) \| f: 19 (36.5%) | m: 84 (81.6%) \| f: 19 (18.4%) | m: 147 (70.0%) \| f: 63 (30.0%) | m: 169 (66.5%) \| f: 85 (33.5%) |
| eGFR [ml/min/1.73m²] | 92.2 (45.6) | 89.8 (47.0) | 71.4 (47.8) | 61.0 (36.7) | 76.6 (51.9) | 88.9 (48.2) |
| Proteinuria [g/24h] | 0.8 (1.2) | 1.1 (1.1) | 1.7 (3.0) | 1.7 (2.6) | 1.4 (2.0) | 0.9 (1.5) |
| MAP [mmHg] | 95.3 (14.8) | 95.0 (16.7) | 95.5 (19.2) | 100.0 (18.7) | 98.3 (18.3) | 95.3 (15.3) |
| M [0\|1] | 0: 155 (76.7%) \| 1: 47 (23.3%) | 0: 67 (62.6%) \| 1: 40 (37.4%) | 0: 34 (65.4%) \| 1: 18 (34.6%) | 0: 60 (58.3%) \| 1: 43 (41.7%) | 0: 127 (60.5%) \| 1: 83 (39.5%) | 0: 189 (74.4%) \| 1: 65 (25.6%) |
| E [0\|1] | 0: 188 (93.1%) \| 1: 14 (6.9%) | 0: 96 (89.7%) \| 1: 11 (10.3%) | 0: 33 (63.5%) \| 1: 19 (36.5%) | 0: 72 (69.9%) \| 1: 31 (30.1%) | 0: 168 (80.0%) \| 1: 42 (20.0%) | 0: 221 (87.0%) \| 1: 33 (13.0%) |
| S [0\|1] | 0: 65 (32.2%) \| 1: 137 (67.8%) | 0: 16 (15.0%) \| 1: 91 (85.0%) | 0: 3 (5.8%) \| 1: 49 (94.2%) | 0: 14 (13.6%) \| 1: 89 (86.4%) | 0: 30 (14.3%) \| 1: 180 (85.7%) | 0: 68 (26.8%) \| 1: 186 (73.2%) |
| T [0\|1\|2] | 0: 170 (84.1%) \| 1: 28 (13.9%) \| 2: 4 (2.0%) | 0: 84 (78.5%) \| 1: 22 (20.6%) \| 2: 1 (0.9%) | 0: 37 (71.2%) \| 1: 15 (28.8%) \| 2: 0 (0.0%) | 0: 78 (75.7%) \| 1: 22 (21.4%) \| 2: 3 (2.9%) | 0: 162 (77.1%) \| 1: 44 (21.0%) \| 2: 4 (1.9%) | 0: 207 (81.5%) \| 1: 43 (16.9%) \| 2: 4 (1.6%) |
| C [0\|1] | 0: 198 (98.0%) \| 1: 4 (2.0%) | 0: 100 (93.5%) \| 1: 7 (6.5%) | 0: 29 (55.8%) \| 1: 23 (44.2%) | 0: 74 (71.8%) \| 1: 29 (28.2%) | 0: 174 (82.9%) \| 1: 36 (17.1%) | 0: 227 (89.4%) \| 1: 27 (10.6%) |

| | | | | | | |
|---|---|---|---|---|---|---|
| Tuft circularity | 0.39 (0.08) | 0.36 (0.08) | 0.31 (0.07) | 0.35 (0.11) | 0.36 (0.09) | 0.37 (0.09) |
| Tuft eccentricity | 0.66 (0.08) | 0.69 (0.11) | 0.72 (0.13) | 0.67 (0.1) | 0.68 (0.1) | 0.67 (0.09) |
| Tuft area [μm²] | 7743.75 (6038.8) | 3021.07 (7340.02) | 2480.11 (5407.37) | 5554.99 (6968.55) | 4333.37 (8029.15) | 7112.28 (8428.37) |
| Tubular diameter [μm] | 28.06 (6.24) | 27.49 (6.1) | 26.71 (7.38) | 28.62 (6.39) | 27.75 (6.56) | 27.73 (6.73) |
| Tubular distance [μm] | 1.97 (1.12) | 2.38 (0.9) | 2.32 (1.1) | 2.26 (1.26) | 2.36 (1.06) | 2.02 (1.01) |
| RASi [y/n] | y: 159 (78.7%) \| n: 43 (21.3%) | y: 77 (72.0%) \| n: 30 (28.0%) | y: 42 (80.8%) \| n: 10 (19.2%) | y: 94 (91.3%) \| n: 9 (8.7%) | y: 171 (81.4%) \| n: 39 (18.6%) | y: 201 (79.1%) \| n: 53 (20.9%) |
| Corticosteroids [y/n] | y: 0 (0.0%) \| n: 202 (100.0%) | y: 107 (100.0%) \| n: 0 (0.0%) | y: 52 (100.0%) \| n: 0 (0.0%) | y: 0 (0.0%) \| n: 103 (100.0%) | y: 107 (51.0%) \| n: 103 (49.0%) | y: 52 (20.5%) \| n: 202 (79.5%) |
| CS therapy duration [years] | 0.0 (0.0) | 1.1 (1.1) | 1.3 (1.5) | 0 (0.0) | 0.5 (1.3) | 0.0 (0.0) |
| Outcome [y/n] | y: 4 (2.0%) \| n: 198 (98.0%) | y: 7 (6.5%) \| n: 100 (93.5%) | y: 3 (5.8%) \| n: 49 (94.2%) | y: 26 (25.2%) \| n: 77 (74.8%) | y: 33 (15.7%) \| n: 177 (84.3%) | y: 7 (2.8%) \| n: 247 (97.2%) |

**Supplementary Table 5.** Validation cohort: characteristics of partitioned subgroups. Continuous variables are reported as median (interquartile range) while categorical variables are reported as absolute (n) and relative (%) frequencies. The primary outcome was defined as a composite of ≥ 50% eGFR reduction and kidney failure within five years of kidney biopsy.

Abbreviations: R, recommended; T, treated; CS, corticosteroid; m, male; f, female; eGFR, estimated glomerular filtration rate; MAP, mean arterial blood pressure; M, mesangial hypercellularity; E, endocapillary hypercellularity; S, segmental glomerulosclerosis; T, tubular atrophy and interstitial fibrosis; C, crescents; RASi, renin-angiotensin-system inhibitor.

| | **R = no CS \| T = no CS** | **R = no CS \| T = CS** | **R = CS \| T = CS** | **R = CS \| T = no CS** | **R ≠ T** | **R = T** |
|---|---|---|---|---|---|---|
| n | 304 | 68 | 44 | 142 | 210 | 348 |
| Follow-up [years] | 5.1 (6.2) | 4.5 (5.6) | 4.8 (7.4) | 4.9 (5.6) | 4.7 (5.6) | 5.1 (6.4) |
| Age [years] | 37.5 (20.0) | 34.2 (22.5) | 30.8 (22.8) | 37.3 (20.6) | 36.5 (21.6) | 36.8 (20.8) |
| Sex [m\|f] | m: 205 (67.4%) \| f: 99 (32.6%) | m: 43 (63.2%) \| f: 25 (36.8%) | m: 27 (61.4%) \| f: 17 (38.6%) | m: 89 (62.7%) \| f: 53 (37.3%) | m: 132 (62.9%) \| f: 78 (37.1%) | m: 232 (66.7%) \| f: 116 (33.3%) |
| eGFR [ml/min²] | 80.5 (46.0) | 86.5 (50.5) | 60.7 (58.3) | 59.9 (50.6) | 72.5 (53.8) | 79.3 (49.2) |
| Proteinuria [g/24h] | 0.9 (1.1) | 1.2 (1.3) | 2.4 (4.1) | 1.6 (3.0) | 1.4 (2.0) | 1.0 (1.2) |
| MAP [mmHg] | 93.3 (17.8) | 95.5 (20.0) | 93.3 (19.6) | 94.8 (16.7) | 94.8 (16.7) | 93.3 (18.0) |
| M [0\|1] | 0: 132 (43.4%) \| 1: 172 (56.6%) | 0: 41 (60.3%) \| 1: 27 (39.7%) | 0: 15 (34.1%) \| 1: 29 (65.9%) | 0: 58 (40.9%) \| 1: 84 (59.2%) | 0: 99 (47.1%) \| 1: 111 (52.9%) | 0: 147 (42.2%) \| 1: 201 (57.8%) |
| E [0\|1] | 0: 260 (85.5%) \| 1: 44 (14.5%) | 0: 56 (82.4%) \| 1: 12 (17.6%) | 0: 27 (61.4%) \| 1: 17 (38.6%) | 0: 81 (57.0%) \| 1: 61 (43.0%) | 0: 137 (65.2%) \| 1: 73 (34.8%) | 0: 287 (82.5%) \| 1: 61 (17.5%) |
| S [0\|1] | 0: 111 (36.5%) \| 1: 193 (63.5%) | 0: 26 (38.2%) \| 1: 42 (61.8%) | 0: 13 (29.6%) \| 1: 31 (70.4%) | 0: 40 (28.2%) \| 1: 102 (71.8%) | 0: 66 (31.4%) \| 1: 144 (68.6%) | 0: 124 (35.6%) \| 1: 224 (64.4%) |
| T [0\|1\|2] | 0: 230 (75.7%) \| 1: 69 (22.7%) \| 2: 5 (1.6%) | 0: 40 (58.8%) \| 1: 26 (38.2%) \| 2: 2 (3.0%) | 0: 20 (45.5%) \| 1: 22 (50.0%) \| 2: 2 (4.5%) | 0: 93 (65.5%) \| 1: 47 (33.1%) \| 2: 2 (1.4%) | 0: 133 (63.3%) \| 1: 73 (34.8%) \| 2: 4 (1.9%) | 0: 250 (71.8%) \| 1: 91 (26.2%) \| 2: 7 (2.0%) |
| C [0\|1] | 0: 285 (93.8%) \| 1: 19 (6.2%) | 0: 58 (85.3%) \| 1: 10 (14.7%) | 0: 22 (50.0%) \| 1: 22 (50.0%) | 0: 76 (53.5%) \| 1: 66 (46.5%) | 0: 134 (63.8%) \| 1: 76 (36.2%) | 0: 307 (88.2%) \| 1: 41 (11.8%) |

| | | | | | | |
|---|---|---|---|---|---|---|
| Tuft circularity | 0.39 (0.08) | 0.35 (0.09) | 0.30 (0.09) | 0.32 (0.09) | 0.33 (0.09) | 0.38 (0.08) |
| Tuft eccentricity | 0.66 (0.09) | 0.66 (0.12) | 0.68 (0.11) | 0.68 (0.13) | 0.68 (0.12) | 0.66 (0.09) |
| Tuft area [µm²] | 7492.56 (9568.65) | 5385.62 (9413.43) | 4358.35 (6479.06) | 2703.81 (6817.52) | 3183.38 (7682.84) | 6824.26 (9510.43) |
| Tubular diameter [µm] | 28.63 (7.16) | 28.59 (6.37) | 29.09 (4.48) | 28.12 (7.09) | 28.33 (6.91) | 28.65 (6.57) |
| Tubular distance [µm] | 2.16 (1.0) | 2.31 (1.21) | 2.68 (1.8) | 2.26 (1.04) | 2.26 (1.08) | 2.26 (1.14) |
| RASi [y/n] | y: 241 (79.3%) \| n: 63 (20.7%) | y: 53 (77.9%) \| n: 15 (22.1%) | y: 36 (81.8%) \| n: 8 (18.2%) | y: 121 (85.2%) \| n: 21 (14.8%) | y: 174 (82.9%) \| n: 36 (17.1%) | y: 277 (79.6%) \| n: 71 (20.4%) |
| Corticosteroids [y/n] | y: 0 (0.0%) \| n: 304 (100.0%) | y: 68 (100.0%) \| n: 0 (0.0%) | y: 44 (100.0%) \| n: 0 (0.0%) | y: 0 (0.0%) \| n: 142 (100.0%) | y: 68 (32.4%) \| n: 142 (67.6%) | y: 44 (12.6%) \| n: 304 (87.4%) |
| CS therapy duration [years] | 0.0 (0.0) | 0.5 (0.7) | 0.8 (0.5) | 0 (0.0) | 0.0 (0.4) | 0.0 (0.0) |
| Outcome [y/n] | y: 18 (5.9%) \| n: 286 (94.1%) | y: 5 (7.4%) \| n: 63 (92.6%) | y: 2 (4.5%) \| n: 42 (95.5%) | y: 24 (16.9%) \| n: 118 (83.1%) | y: 29 (13.8%) \| n: 181 (86.2%) | y: 20 (5.7%) \| n: 328 (94.3%) |

**Supplementary Table 6.** Scoring of tubulointerstitial inflammation. 60 cases with increased tubular distance (30 with high predicted benefit suggestive for active tubulointerstitial inflammation and 30 with low predicted benefit suggestive for tubulointerstitial scarring) were scored blinded in accordance with the Banff classification for kidney transplant pathology.
Abbreviations: ITE, individualized treatment effect; i, interstitial inflammation; t, tubulitis.

| | **Low predicted ITE** | **High predicted ITE** |
|---|---|---|
| Overall n | 30 | 30 |
| **Interstitial inflammation (i)** | | |
| i0 | 27 (90.0%) | 18 (60.0%) |
| i1 | 2 (6.67%) | 7 (23.33) |
| i2 | 1 (3.33%) | 3 (10.0%) |
| i3 | 0 (0%) | 2 (6.67%) |
| **Tubulitis (t)** | | |
| t0 | 25 (83.33%) | 16 (53.33%) |
| t1 | 5 (16.67%) | 13 (43.33%) |
| t2 | 0 (0%) | 0 (0%) |
| t3 | 0 (0% | 1 (3.33%) |

**Supplementary Table 7.** Robustness analyses comparing the predicted average treatment effect of the fitted model to random perturbations of covariates, treatment and outcome.

| | |
|---|---|
| **Complete model** | |
| Average treatment effect | 0.254 (95% CI 0.244-0.268) |
| **Random predictor** | |
| Average treatment effect | 0.236 (95% CI 0.224-0.25) |
| **Random replace** | |
| Average treatment effect | 0.227 (95% CI 0.209-0.251) |
| **Random treatment** | |
| Average treatment effect | -0.05 (95% CI -0.071-(-0.029)) |
| **Random outcome** | |
| Average treatment effect | 0.094 (95% CI 0.068-0.119) |

**Supplementary Table 8.** TRIPOD+AI checklist.

| Section/Topic | Item | Development /Validation | Checklist item | Reported on page |
|---|---|---|---|---|
| **Title** | | | | |
| Title | 1 | D; V | Identify the study as developing or evaluating the performance of a multivariate prediction model, the target population, and the outcome to be predicted | p. 2 |
| **Abstract** | | | | |
| Abstract | 2 | D; V | See TRIPOD+AI for Abstracts checklist | p. 2 |
| **Introduction** | | | | |
| Background | 3a | D; V | Explain the healthcare context (including whether diagnostic or prognostic) and rationale for developing or evaluating the prediction model, including references to existing models | p. 3 |
| | 3b | D; V | Describe the target population and the intended purpose of the prediction model in the context of the care pathway, including its intended users (e.g., healthcare professionals, patients, public) | p. 3 |
| | 3c | D; V | Describe any known health inequalities between sociodemographic groups | p. 3 |
| Objectives | 4 | D; V | Specify the study objectives, including whether the study describes the development or validation of a prediction model (or both) | p. 4 |
| **Methods** | | | | |
| Data | 5a | D; V | Describe the sources of data separately for the development and validation datasets (e.g., randomised trial, cohort, routine care or registry data), the rationale for using these data, and representativeness of the data | p. 10 |
| | 5b | D; V | Specify the dates of the collected participant data, including start and end of participant accrual; and, if applicable, end of follow-up | Supp. p. 14-15 |
| Participants | 6a | D; V | Specify key elements of the study setting (e.g., primary care, secondary care, general population) including the number and location of centres | Supp. p. 14-15 |
| | 6b | D; V | Describe the eligibility criteria for study participants | p. 10-11. Fig. 1, Supp. p. 14 |
| | 6c | D; V | Give details of any treatments received, and how they were handled during model development or evaluation, if relevant | p. 10-11. Fig. 1, Supp. p. 14 |
| Data preparation | 7 | D; V | Describe any data pre-processing and quality checking, including whether this was similar across relevant sociodemographic groups | p. 11, Supp. p. 15-17 |
| Outcome | 8a | D; V | Clearly define the outcome that is being predicted and the time horizon, including how and when assessed, the rationale for choosing this outcome, and whether the method of outcome assessment is consistent across sociodemographic groups | p. 11-12 |
| | 8b | D; V | If outcome assessment requires subjective interpretation, describe the qualifications and demographic characteristics of the outcome assessors | NA |
| | 8c | D; V | Report any actions to blind assessment of the outcome to be predicted | Supp. p. 19-20 |
| Predictors | 9a | D | Describe the choice of initial predictors (e.g., literature, previous models, all available predictors) and any pre-selection of predictors before model building | p. 11, Supp. p. 15-16 |

| | | | | |
|---|---|---|---|---|
| | 9b | D; V | Clearly define all predictors, including how and when they are measured (and any actions to blind assessment of predictors for the outcome and other predictors) | p. 11, Supp. p. 15-16 |
| | 9c | D; V | If predictor measurement requires subjective interpretation, describe the qualifications and demographic characteristics of the predictor assessors | Supp. p. 15-16 |
| Sample size | 10 | D; V | Explain how the study size was arrived at (separately for the development and evaluation) and justify that the study size was sufficient to answer the research question. Include details of any sample size calculation | p. 4, Supp. Figure 1 |
| Missing data | 11 | D; V | Describe how missing data were handled. Provide reasons for omitting any data | Supp. p. 16 |
| Analytical methods | 12a | D | Describe how the data were used (e.g., for development and evaluation of model performance) in the analysis, including whether the data were partitioned, considering any sample size requirements | p. 10-12 |
| | 12b | D | Depending on the type of model, describe how predictors were handled in the analyses (functional form, rescaling, transformation, or any standardization) | p. 11, Supp. p. 15-16 |
| | 12c | D | Specify the type of model, rationale, all model-building steps, including any hyperparameter tuning, and method for internal validation | p. 11-12, Supp. 17-18 |
| | 12d | D; V | Describe if and how any heterogeneity in estimates of model parameter values and model performance was handled and quantified across clusters (e.g., hospitals, countries). See TRIPOD-Cluster for additional considerations. | p. 12, Supp. p. 19 |
| | 12e | D; V | Specify all measures and plots used (and their rationale) to evaluate model performance (e.g., discrimination, calibration, clinical utility) and, if relevant, to compare multiple models | p. 11-12, Supp. p. 18 |
| | 12f | V | Describe any model updating (e.g., recalibration) arising from the model evaluation, either overall or for any particular sociodemographic groups or settings | NA |
| | 12g | V | For model evaluation, describe how the model predictions were calculated (e.g., formula, code, object, application programming interface) | p. 11-12, Supp. p. 18 |
| Class imbalance | 13 | D; V | If class imbalance methods were used, state why and how this was done, and any subsequent methods to recalibrate the model or the model predictions | p. 11, Supp. p. 17-18 |
| Fairness | 14 | D; V | Describe any approaches that were used to address model fairness and their rationale | Supp. p. 14-15; 19 |
| Model output | 15 | D | Specify the output of the prediction model (e.g., probabilities, classification). Provide details and rationale for any classification and how the thresholds were identified | p. 11-12, Supp. p. 18 |
| Training versus evaluation | 16 | D; V | Identify any differences between the development and evaluation data in healthcare setting, eligibility criteria, outcome and predictors | p. 10-11, Supp. p. 14-16 |
| Ethical approval | 17 | D; V | Name the institutional research board or ethics committee that approved the study and describe the participant-informed consent or the ethics committee waiver of informed consent | p. 13 |
| **Open Science** | | | | |
| Funding | 18a | D; V | Give the source of funding and the role of the funders for the present study | p. 13-14 |
| Conflict of interest | 18b | D; V | Declare any conflicts of interest and financial disclosures for all authors | p. 13 |
| Protocol | 18c | D; V | Indicate where the study protocol can be accessed or state that a protocol was not prepared | p. 12-13 |
| Registration | 18d | D; V | Provide registration information for the study, including register name and registration number, or state that the study was not registered | p. 14, Supp. p. 4-13 |

| | | | | |
|---|---|---|---|---|
| Data sharing | 18e | D; V | Provide details of the availability of the study data | p. 12-13 |
| Code sharing | 18f | D; V | Provide details of the availability of the analytical code | p. 12-13 |
| **Patient & Public involvement** | | | | |
| Patient & Public Involvement | 19 | D; V | Provide details of any patient and public involvement during the design, conduct, reporting, interpretation, or dissemination of the study or state no involvement. | NA |
| **Results** | | | | |
| Participants | 20a | D; V | Describe the flow of participants through the study, including the number of participants with and without the outcome and, if applicable, a summary of the follow-up time. | Figure 1, Table 1, p. 4 |
| | 20b | D; V | Report the characteristics overall and, where applicable, for each data source or setting, including the key dates, key predictors (including demographics), treatments received, sample size, number of outcome events, follow-up time, and amount of missing data. Report any differences across key demographic groups. | Table 1, Supp. Table 1-2 |
| | 20c | V | For model evaluation, show a comparison with the development data of the distribution of important predictors (demographics, predictors, and outcome) | Table 1, Supp. Table 1-2 |
| Model development | 21 | D; V | Specify the number of participants and outcome events in each analysis (e.g., for model development, hyperparameter tuning, model evaluation) | Table 1, Supp. Table 1-2 |
| Model specification | 22 | D | Provide details of the full prediction model (e.g., formula, code, object, application programming interface) to allow predictions in new individuals and to enable third-party evaluation and implementation, including any restrictions to access or re-use (e.g., freely available, proprietary) | p. 12-13 |
| Model performance | 23a | D; V | Report model performance estimates with confidence intervals, including for any key subgroups (e.g., sociodemographic). | p. 5-6 |
| | 23b | D; V | If examined, report results of any heterogeneity of model performance across clusters. See TRIPOD-Cluster for additional details. | NA |
| Model updating | 24 | V | Report the results from any model updating, including the updated model and subsequent performance. | NA |
| **Discussion** | | | | |
| Interpretation | 25 | D; V | Give an overall interpretation of the main results, including issues of fairness in the context of the objectives and previous studies | p. 8 |
| Limitations | 26 | D; V | Discuss any limitations of the study (such as non-representative sample, sample size, overfitting, missing data) and their effects of any biases, statistical uncertainty, and generalizability | p. 9 |
| Usability of the model in the context of current care | 27a | D | Describe how poor quality or unavailable input data (e.g., predictor values) should be assessed and handled when implementing the prediction model | p. 9 |
| | 27b | D | Specify whether users will be required to interact in the handling of the input data or use of the model, and what level of expertise is required of users | p. 9 |
| | 27c | D; V | Discuss any next steps for future research, with a specific view to applicability and generalizability of the model | p. 9 |

**Supplementary Table 9.** Definitions for the five included pathomics features.

| **Pathomics predictor** | **Definition** |
|---|---|
| Tuft circularity | Shape of the segmented glomerular tuft, measuring how circular the structure is: $C = \frac{(4* \pi * Area)}{Perimeter^2}$. The circularity of a perfect circle equals 1. |
| Tuft eccentricity | Shape of the segmented glomerular tuft, measuring the ratio of axes of the best-fit ellipse to the tuft: $Ecc = \frac{\sqrt{(major\ axis\ length^2 - minor\ axis\ length^2)}}{major\_axis\_length}$. The eccentricity of a perfect circle equals 0 and increases with greater elongation. |
| Tuft area | Area of the segmented glomerular tuft section [μm²]. |
| Tubular diameter | Diameter of the largest circle fully fitting inside the segmented tubular instance [μm]. |
| Tubular distance | Closest distance between the borders of a segmented tubule to its neighboring instances [μm]. |

**Supplementary Table 10.** Overview of datasets, study groups and contributors**.**

| **Dataset** | **Contact** | **Contact** |
|---|---|---|
| VALIGA | Rosanna Coppo for the VALIGA investigators | |
| Kyoto | Motoko Yanagita | |
| NURTuRE-CKD | Maarten W. Taal & Philip A. Kalra for the NURTuRE academic steering group | |
| Aachen | Peter Boor | |
| Leicester | Jonathan Barratt | |
| Rochester | Andrew Rule | |
| Diyarbakir | Ulaş Alabalik | |
| CureGN | Laura Barisoni for the CureGN investigators | Data are available upon request through the CureGN Ancillary Studies program. Data access is governed by the CureGN Steering Committee and NIDDK. Additional data sets will be provided to the NIDDK Central Repository after completion of study recruitment, which is currently ongoing. After data are deposited, the data will be available through the NIDDK Central Repository (https://repository.niddk.nih.gov/home/). |

**Supplementary Table 11.** Missingness of patient characteristics in derivation and validation cohort in absolute (n) and relative (%) frequencies.
Abbreviations: M, mesangial hypercellularity; E, endocapillary hypercellularity; S, segmental glomerulosclerosis; T, tubular atrophy and interstitial fibrosis; C, crescents; RASi, renin-angiotensin-system inhibitor; CS, corticosteroid.

| | **Derivation** | **Validation** |
|---|---|---|
| n | 464 | 558 |
| Follow-up | 0 (0%) | 0 (0%) |
| Age | 0 (0%) | 0 (0%) |
| Sex | 0 0%) | 0 0%) |
| Race and ethnicity | 5 (1.08%) | 68 (12.19%) |
| BMI | 64 (13.79%) | 189 (33.87%) |
| eGFR | 0 (0%) | 0 (0%) |
| Proteinuria | 14 (3.02%) | 35 (6.27%) |
| MAP | 2 (0.43%) | 163 (29.21%) |
| M | 0 (0%) | 0 (0%) |
| E | 0 (0%) | 0 (0%) |
| S | 0 0%) | 0 0%) |
| T | 0 (0%) | 0 (0%) |
| C | 0 (0%) | 0 (0%) |
| Tuft circularity | 0 (0%) | 0 (0%) |
| Tuft eccentricity | 0 (0%) | 0 (0%) |
| Tuft area | 0 0%) | 0 0%) |
| Tubular Diameter | 0 (0%) | 0 (0%) |
| Tubular Distance | 0 (0%) | 0 (0%) |
| RASi | 0 (0%) | 0 (0%) |
| CS | 0 (0%) | 0 (0%) |
| CS therapy duration | 20 (4.31%) | 140 (25.09%) |
| Outcome | 0 (0%) | 0 (0%) |

**Supplementary Figure 1.** Baseline cumulative event probability (A) and overall treatment effect (B).

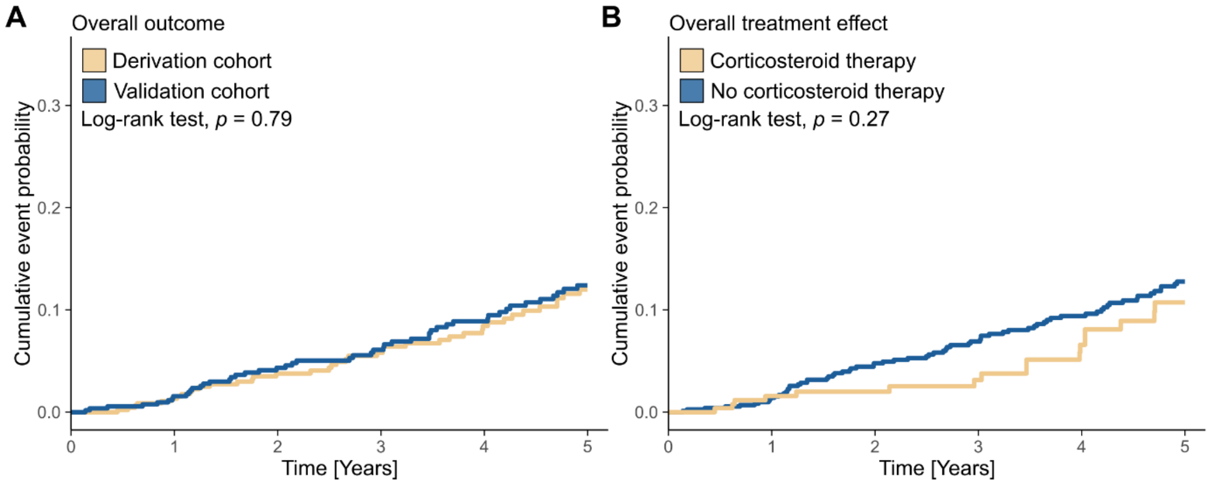

**Supplementary Figure 2.** Qini (A) and rank-weighted average treatment effect (B) for the Xboost model in the derivation cohort. Patients were ranked based on their predicted individualized treatment benefit from most to least likely to benefit from corticosteroid treatment. The straight dotted line in (A) represents a random treatment allocation which is compared to the model's predicted uplift. The computed Qini coefficient is the area between the solid curve and dotted line. Based on the increase in benefit for the grouped quantiles, an individualized treatment policy was derived in derivation and validation cohorts.

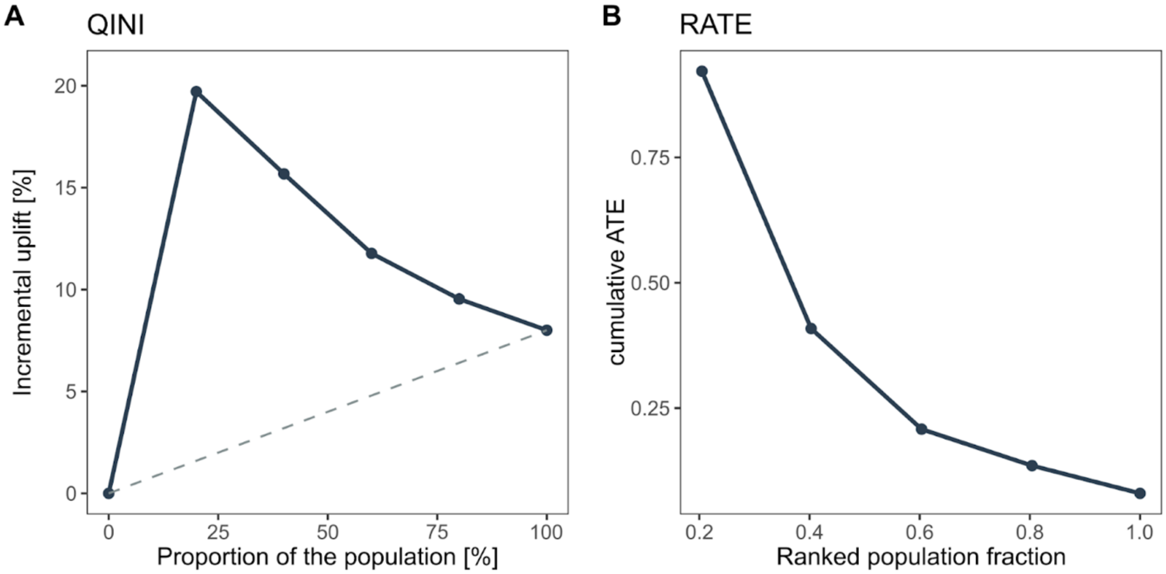

**Supplementary Figure 3.** Cumulative event probability for treatment recommendations in the derivation and validation cohorts. Model-based recommendations were compared with the observed treatment resulting in four possible scenarios (A-B). Out of these four recommendations, patients could be further stratified into whether they received the model-recommended treatment or not (C-D). (D) shows the cumulative event probability at five years in the derivation cohort for recommended versus non-recommended treatment.

Abbreviations: R, recommendation; T, treatment; CS, corticosteroids.

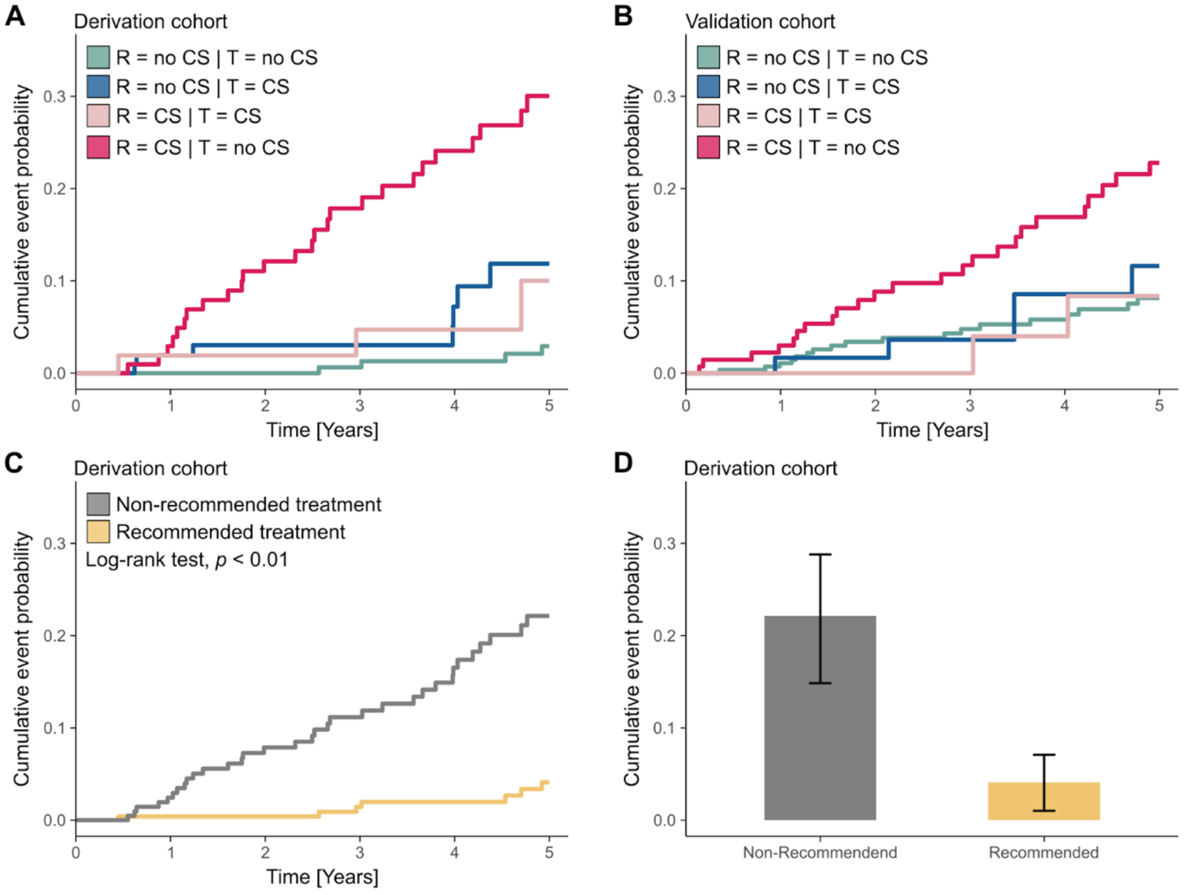

**Supplementary Figure 4.** Calibration of the Xboost model in the validation cohort. Calibration was assessed by comparing group-level predictions of treatment effect with the observed treatment effect of each group. Both predicted and observed benefit was normalized for visualization.

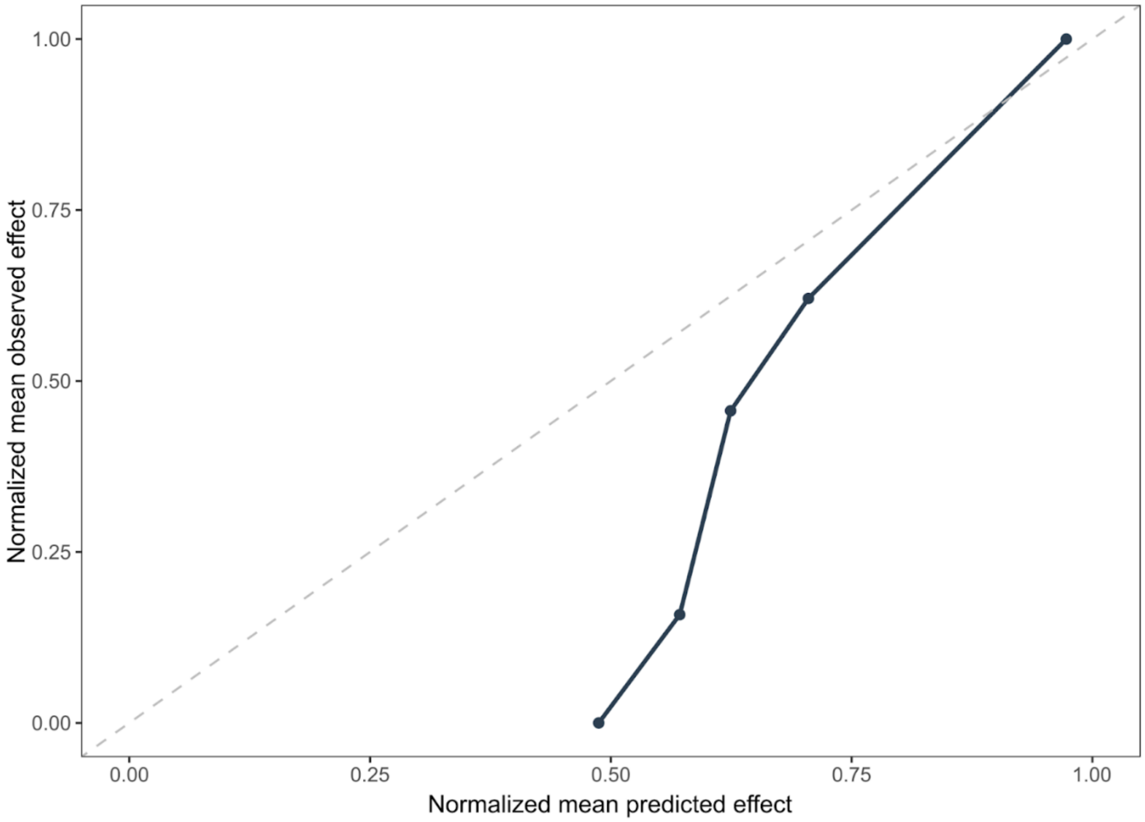

**Supplementary Figure 5.** Shapley additive explanations (SHAP) values of representative cases from the validation cohort with high (A-B) and low (C) predicted treatment benefit. The x-axis includes the six most important predictors which were sequentially added to the prediction to demonstrate the change from average treatment effect (ATE) to individualized treatment effect (ITE).

Abbreviations: gfr, estimated glomerular filtration rate; E, endocapillary hypercellularity; C, crescents.

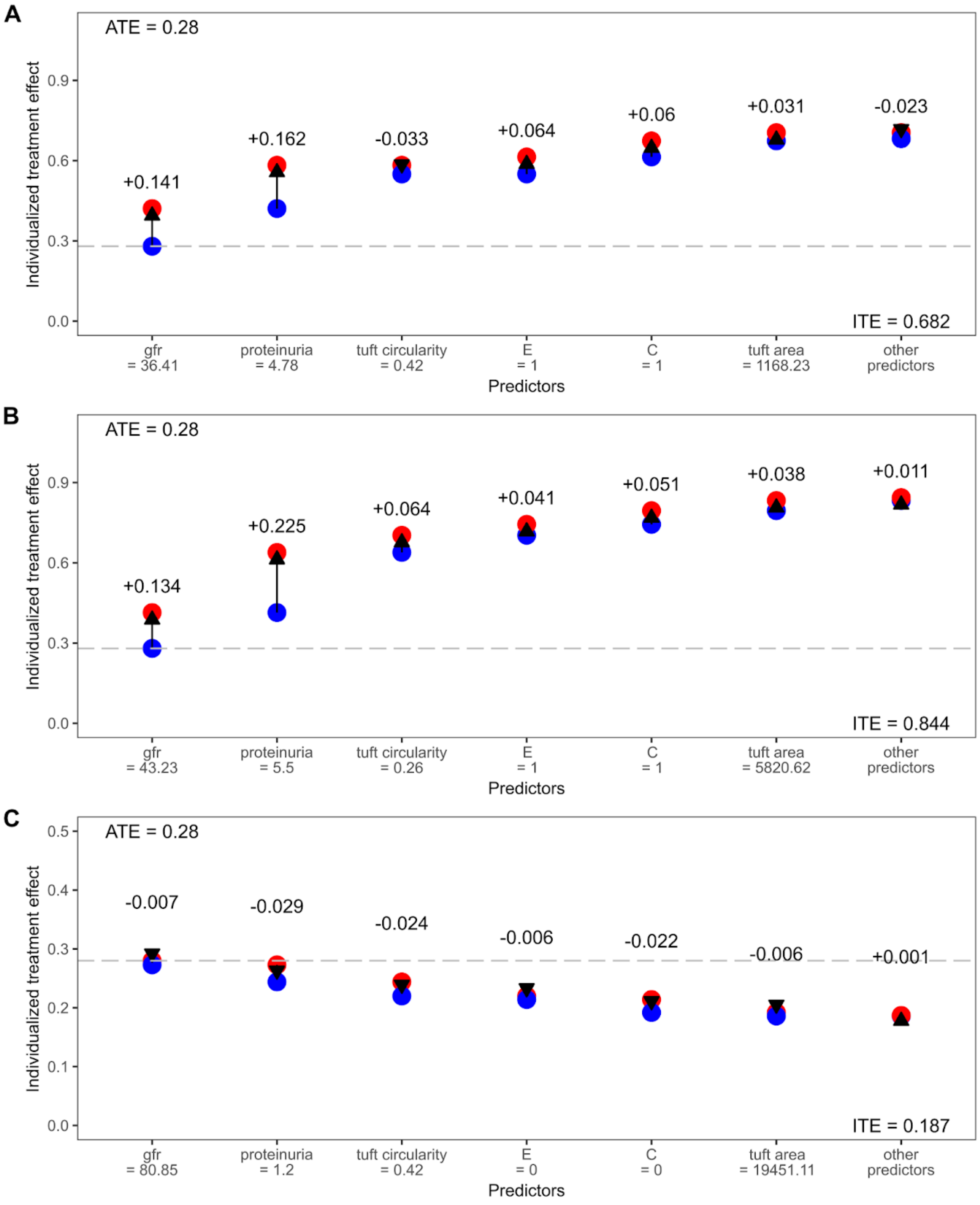

**Supplementary Figure 6.** Shapley additive explanations (SHAP) values of MEST-C predictors (A-E) calculated in the validation cohort.

Abbreviations: M, mesangial hypercellularity; E, endocapillary hypercellularity; S, segmental glomerulosclerosis; T, tubular atrophy and interstitial fibrosis; C, crescents.

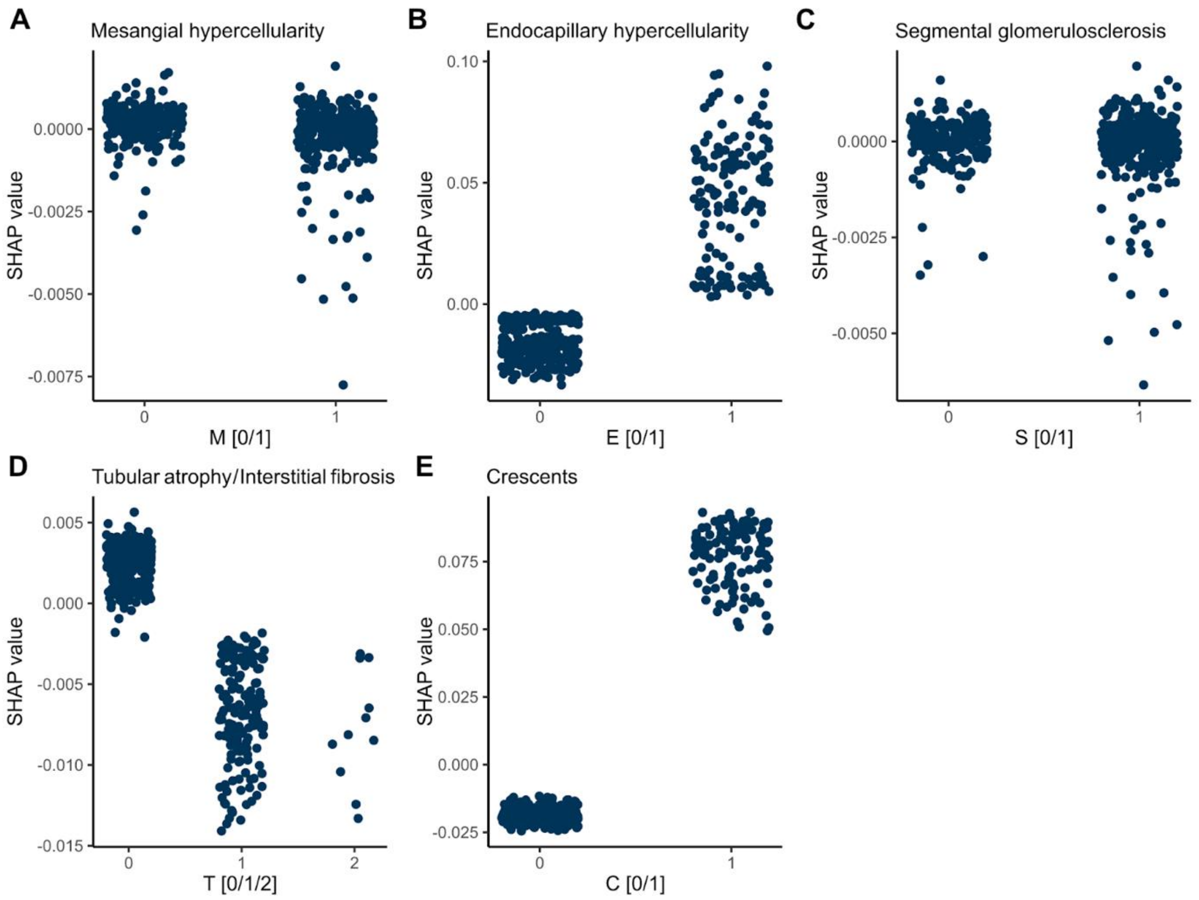

**Supplementary Figure 7.** Representative visualizations of glomeruli in patients with high (A) or low predicted treatment benefit (B) based on the associated glomerular tuft circularity. All images represent patches of 500x500µm.

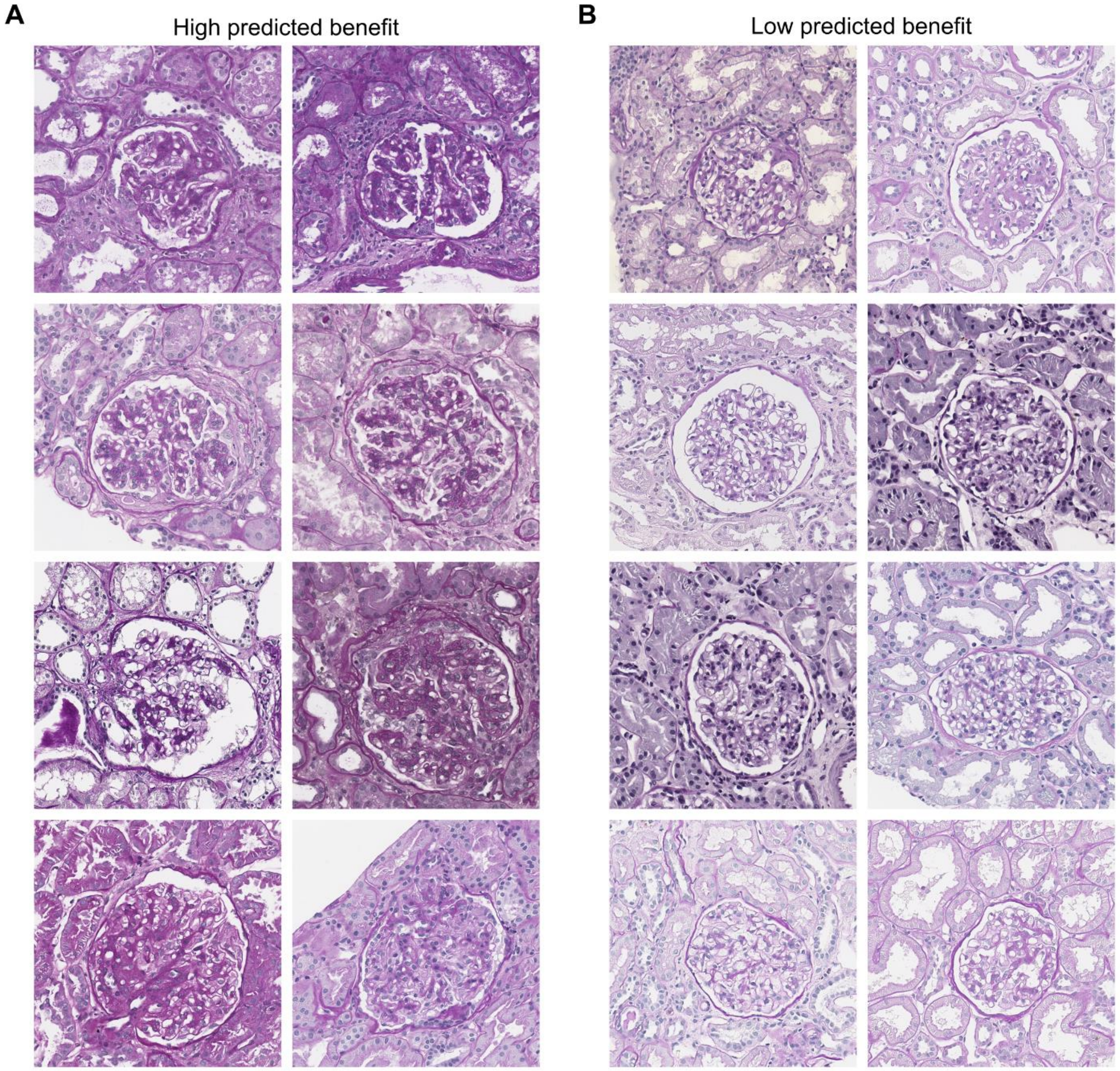

**Supplementary Figure 8.** Distribution of predicted individualized treatment effects in both derivation and validation cohort (A), as well as in all subcohorts (B).

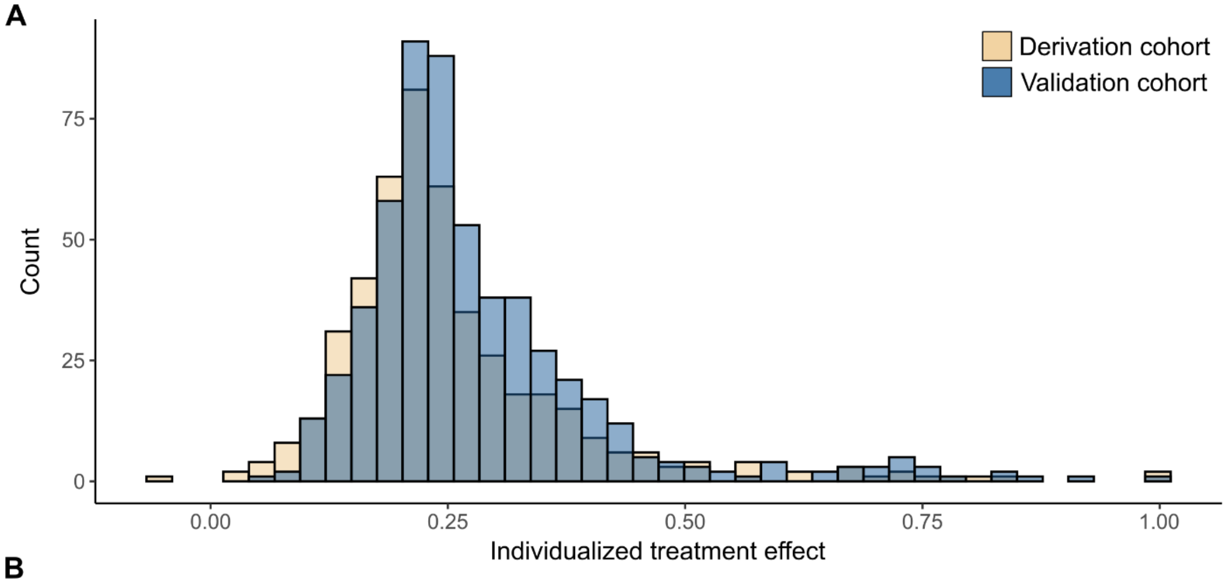


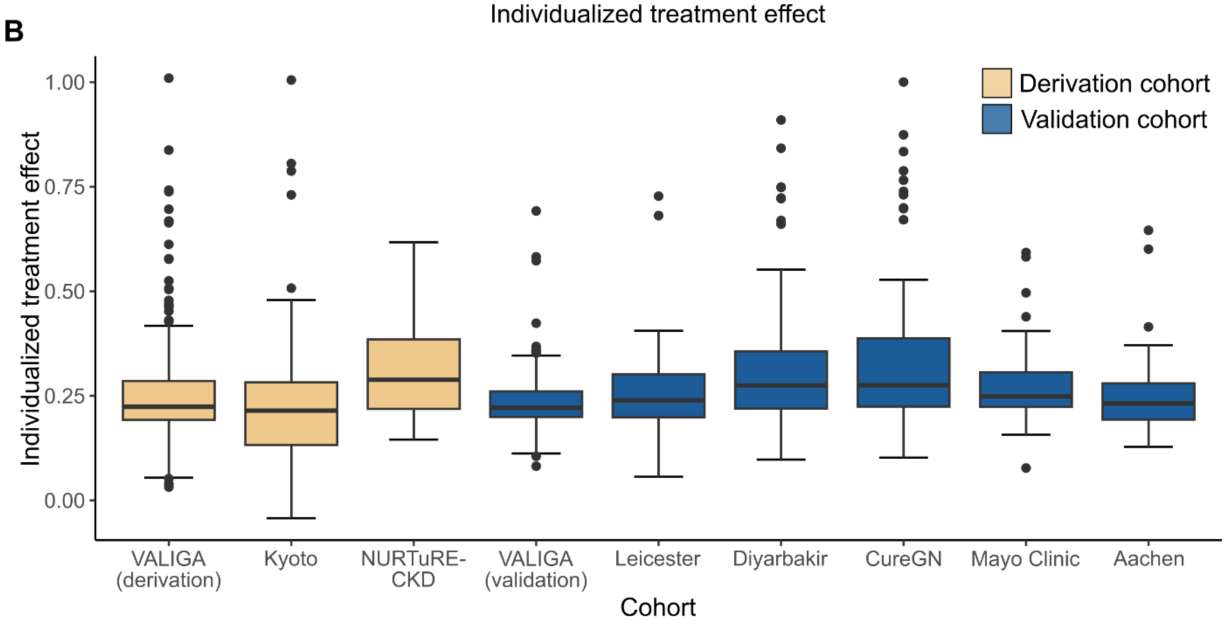
